# Repetitive Transcranial Magnetic Stimulation Modulates Brain Connectivity in Children with Self-limited Epilepsy with Centrotemporal Spikes

**DOI:** 10.1101/2024.08.27.24312648

**Authors:** Xiwei She, Wendy Qi, Kerry C. Nix, Miguel Menchaca, Christopher C. Cline, Wei Wu, Zihuai He, Fiona M. Baumer

## Abstract

**Objective:** Interictal epileptiform discharges (IEDs) alter brain connectivity in children with epilepsy; this connectivity change may be a mechanism by which epilepsy induces cognitive deficits. Here, we test whether repetitive transcranial magnetic stimulation (rTMS), a non-invasive neuromodulation technique, modulates connectivity and reduces IEDs in children with epilepsy.

**Methods:** Nineteen children with self-limited epilepsy with centrotemporal spikes (SeLECTS) participated in a cross-over study comparing the impact of active vs. sham rTMS on IEDs and brain connectivity. SeLECTS is an epilepsy syndrome affecting the motor cortex, and prior studies show that motor cortices become pathologically hyper-connected to frontal and temporal language cortices. Using a crossover design, we compared the effect of single doses of active versus sham motor cortex rTMS. Connectivity, which was quantified by the weighted phase lag index (wPLI), was measured before and after rTMS using single pulses of TMS combined with EEG (spTMS-EEG). Analyses focused on six regions: bilateral motor cortices and bilateral inferior frontal and superior temporal regions. IEDs were counted in the five minutes before and after rTMS.

**Results:** Active, but not sham, rTMS significantly and globally decreased wPLI connectivity between multiple regions, with the greatest reductions seen in the superior temporal region connections in the stimulated hemisphere. Additionally, there was a trend suggesting that rTMS decreases IED frequency.

**Interpretation:** These findings underscore the potential of low-frequency rTMS to target pathologic hyperconnectivity and reduce IEDs in children with SeLECTS and potentially other pediatric epilepsy syndromes, offering a promising avenue for therapeutic intervention.

## Introduction

Interictal epileptiform discharges (IEDs) are brief bursts of abnormal hypersynchronous cortical activity occurring in patients with epilepsy. Children with epilepsy who experience frequent IEDs often suffer cognitive deficits including language deficits,^1^ but how IEDs cause these deficits is not fully understood. Prior studies suggest that IEDs induce aberrant hyperconnectivity between the epileptic cortex, where IEDs originate, and distant brain regions, including key nodes in the language network.^2,3^ This hyperconnectivity persists even during IED-free period,^4^ and is associated with worse language performance.^5^ Therapies aimed at reducing hyperconnectivity may therefore alleviate cognitive deficits in children with epilepsy.

Transcranial magnetic stimulation (TMS) combined with electroencephalography (EEG) is a powerful tool for investigating and modulating the neurophysiological properties of the brain.^6,7^ Single pulses of TMS activate specific brain regions, while EEG (spTMS-EEG) captures the resultant network responses as TMS evoked potentials (TEPs). TEPs can be measured locally and globally to assess cortical excitability and connectivity, respectively.^8,9^ In addition to measure evoked responses in time domain, connectivity can also be investigated by analyzing the relationship of inter-regional evoked responses using phase synchronization measurements.^10,11^

When TMS is applied in repetitive patterns (rTMS), it modulates brain activity for a period exceeding the stimulation train. Studies of healthy adults show that rTMS can suppress or facilitate cortical excitability depending upon the stimulation pattern,^12^ with low-frequency stimulation (≤1 Hz) generally reducing excitability. Furthermore, rTMS can affect excitability of functionally connected brain regions,^13^ and several sham-controlled trials in adults with epilepsy suggest that rTMS suppresses IEDs.^14,15^ Controlled studies in pediatric epilepsy are still lacking.

This study investigates the impact of 1 Hz rTMS on cortical excitability, connectivity, and IED frequency using spTMS-EEG in 19 children with self-limited epilepsy with centrotemporal spikes (SeLECTS). SeLECTS provides a unique model for isolating the effect of IEDs on cognition, because children have rare seizures and are generally otherwise healthy; however, they have mild to moderate difficulties with language and frequent IEDs from the central motor regions.^16^ Using a crossover design, we delivered active or sham 1 Hz rTMS to the motor cortex with the highest frequency of IEDs. Single-pulse TMS was applied to the same motor cortex, with spTMS-EEG analyses focused on six regions of interest (ROIs): the stimulated and contralateral motor cortices (implicated in the epilepsy), as well as the bilateral inferior frontal and superior temporal regions (important for language).^17^ We investigated three measurements: 1) TEPs measured at the stimulated motor cortex and at the other five ROIs, providing a time-domain assessment of cortical excitability and connectivity, respectively; 2) the weighted phase lag index (wPLI),^10^ a phase-domain connectivity measure that is robust against the confounding effects of volume conduction that can obscure temporal domain connectivity analysis, between all ROIs; and 3) frequency of IEDs. We aim to determine if 1 Hz rTMS modulates aberrant hyperconnectivity and IEDs, offering a step toward developing novel epilepsy treatments.

## Methods

### Participants

Children between the ages of 5–18 years with a diagnosis of SeLECTS based on seizure semiology (hypersalivation, facial/hemibody twitching, or nocturnal tonic-clonic) and centrotemporal IEDs were eligble.^16^ We excluded children with a history of: prematurity (<35 weeks); serious neurologic problems (e.g., head trauma, cerebrovascular accident or neuro-inflammatory disease); or focal deficits on neurologic exam. Children were recruited from Lucile Packard Children’s Hospital. This study was approved by the Stanford University Institutional Review Board. Written consent was obtained from parents and assent from children. Antiseizure medication (ASM) use and serum level were recorded.

### TMS Experiment

#### *Overview* (Fig. 1A)

The study was conducted over two sessions separated by a minimum of 6 days to allow for wash-out of rTMS effects. Procedures were identical except that during one session, children underwent active rTMS while during the other they underwent sham rTMS. Session order was randomized. Sessions were conducted in the late morning or early afternoon, with participants comfortably seated in a semi-reclined position.

**Figure 1.**
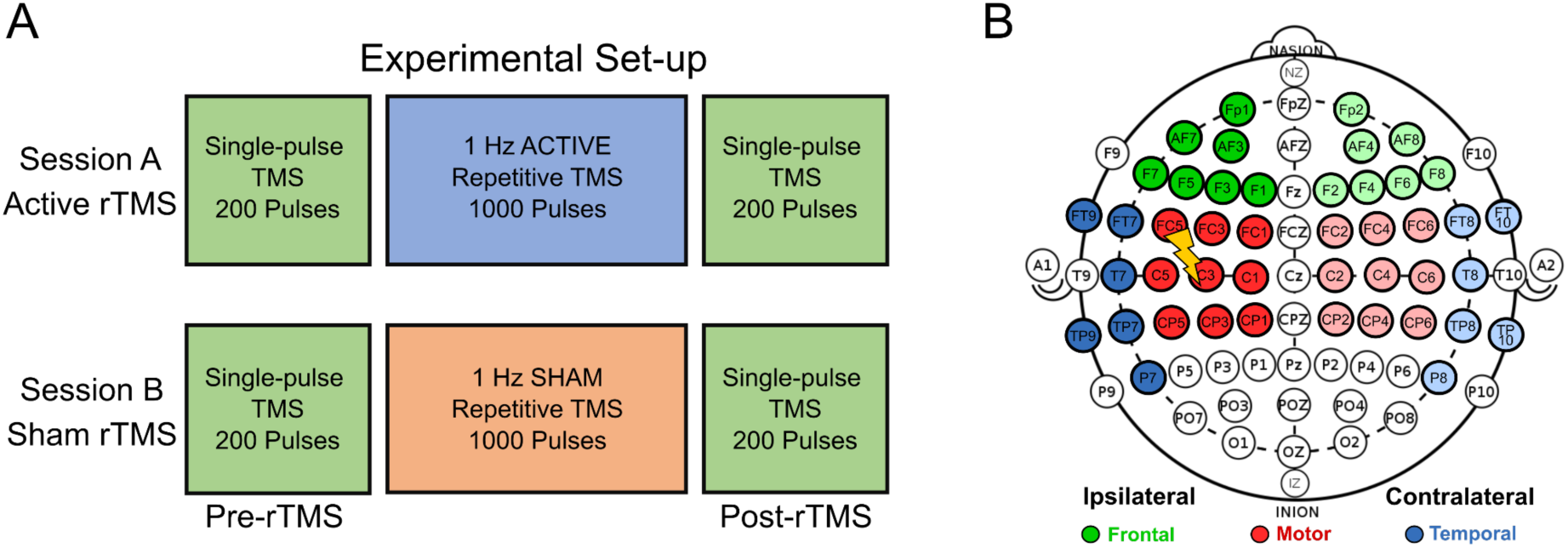
TMS and EEG Data Collection. A: Experimental protocol with participants undergoing spTMS-EEG to the motor cortex (green boxes) before and after either active (blue) or sham (orange) rTMS. Session order was randomized on a per-participant basis. B: High-density EEG was recorded. The hemisphere to which rTMS was applied is called “ipsilateral” and the opposite hemisphere “contralateral.” Regions of interest included: ipsilateral and contralateral frontal (green), motor (red), and temporal (blue).

#### EEG Recordings

EEG recordings were acquired using a BrainVision actiCHamp Pulse amplifier with 64-channel actiCAP with slim active electrodes, at a sampling rate of 25 kHz. Impedances were kept below 10 kΩ to balance feasible setup time for children with adequate data quality.^18,19^

#### TMS

TMS was administered using a MagVenture MagPro X100 stimulator equipped with a Cool-B65 figure-8 coil, and guided by the Localite TMS Navigator neuro-navigation system, registered to a representative magnetic resonance image (MRI) from unbiased average age-appropriate templates.^20^ Electromyographic (EMG) were measured from the bilateral abductor pollicis brevis (APB) muscles. The motor hotspot was identified as the location eliciting the largest EMG deflection in the APB EMG electrode when the hand was relaxed. On both days, the resting motor threshold (rMT) was determined as the minimum intensity required to evoke a peak-to-peak EMG signal of at least 50 µV in at least 5 out of 10 pulses.^21^ Younger children and people taking ASMs have elevated motor thresholds.^19,22–24^ If no EMG deflection was observed at maximal stimulator output (MSO), hot-spotting was performed with the hand slightly contracted and the rMT was defined as 100% MSO. To minimize auditory contamination, in-ear headphones played noise matched in frequency to the TMS clicks at a volume at which children reported difficulty hearing clicks but experienced no discomfort.^25^

We additionally applied a foam layer to the coil surface to reduce TMS-induced vibration. Alertness was confirmed and maintained throughout sessions via monitoring patient behavior and EEG.

#### Repetitive TMS

We targeted rTMS to the hand motor hotspot of the hemisphere that had the greatest frequency of IEDs on most recent clinical EEG. We chose this location as seizures and IEDs in SeLECTS originate in the motor cortex and TMS to the hand region of motor cortex is tolerable. We administered 1000 pulses of 1 Hz active or sham rTMS, whose intensity was 90% of determined rMT. Active and sham sessions were separated by at least 6 days, with condition order randomized on a per-participant basis (Fig. 1A).

#### Single-Pulse TMS

Before and after the rTMS, participants underwent 200-pulse blocks of spTMS-EEG to the same motor hotspot of rTMS administration. Stimulation intensity was 120% rMT (or 100% MSO if rMT exceeded 84% MSO), with pulses jittered at random intervals of 2-3 seconds. The spTMS-EEG was used to quantify the impact of rTMS.

### Data Analyses

#### spTMS-EEG Data Preprocessing

We preprocessed the pre- and post-rTMS spTMS–EEG data using a EEGLAB-based,^26^ custom-designed spTMS-EEG preprocessing pipeline.^19,27^ Data were epoched from −1000 to 1500 milliseconds around the spTMS pulse. To eliminate the spTMS pulse artifact, an interpolation procedure was applied −2 to 12 milliseconds relative to pulse onset. Data were down-sampled to 1 kHz, baseline-corrected by subtracting the mean within the −500 to −10 millisecond timespan from all data points, and high-pass filtered at 1 Hz.

Bad channel rejection was performed using a data-driven Wiener noise estimation method.^28^ Further noise reduction utilized the SOUND algorithm,^29^ and line noise was attenuated by a Butterworth band stop filter (58-62 Hz). Independent component analysis, via the ICLabel algorithm,^30^ identified and eliminated artifacts. Following the ICLabel step, components were screened for remaining artifacts during the 11 to 30-millisecond time window, where TMS-induced muscle artifacts typically occur. This rejection rule was inspired by a similar approach from the TESA toolbox.^31^ The signal was then low-pass filtered below 200 Hz and re-referenced to a common average. After these steps, trials with prominent muscle artifacts were rejected through visual inspection and was confirmed by a pediatric epileptologist (FMB).

#### Regions of Interest (ROIs)

We categorized the hemispheres as either ipsilateral or contralateral to the rTMS administration (Fig. 1B). We focused on six ROIs: the ipsilateral and contralateral frontal, motor, and temporal regions. We chose these regions as IEDs emerge from the motor cortex in SeLECTS and frontal and temporal regions are important for language function.^17^

#### TMS Evoked Potentials (TEPs; Fig. 2A)

TEPs reflect cortical reactivity and propagation along functional pathways.^6,9^ **Local TEPs** can be measured at the site of stimulation directly under the coil, where they reflect a mixture of local and long-range excitatory and inhibitory inputs.^8^ We used the local TEP as a *measurement of cortical excitability*. **Remote TEPs** measured in brain regions distant from the stimulated cortex were used as a *measure of connectivity* between the stimulated and recorded regions.^9^ For both TEP measurements, we first averaged EEG signals from multiple spTMS epochs to create a “grand average” TEP from which we derived the following measurements:

**Figure 2.**
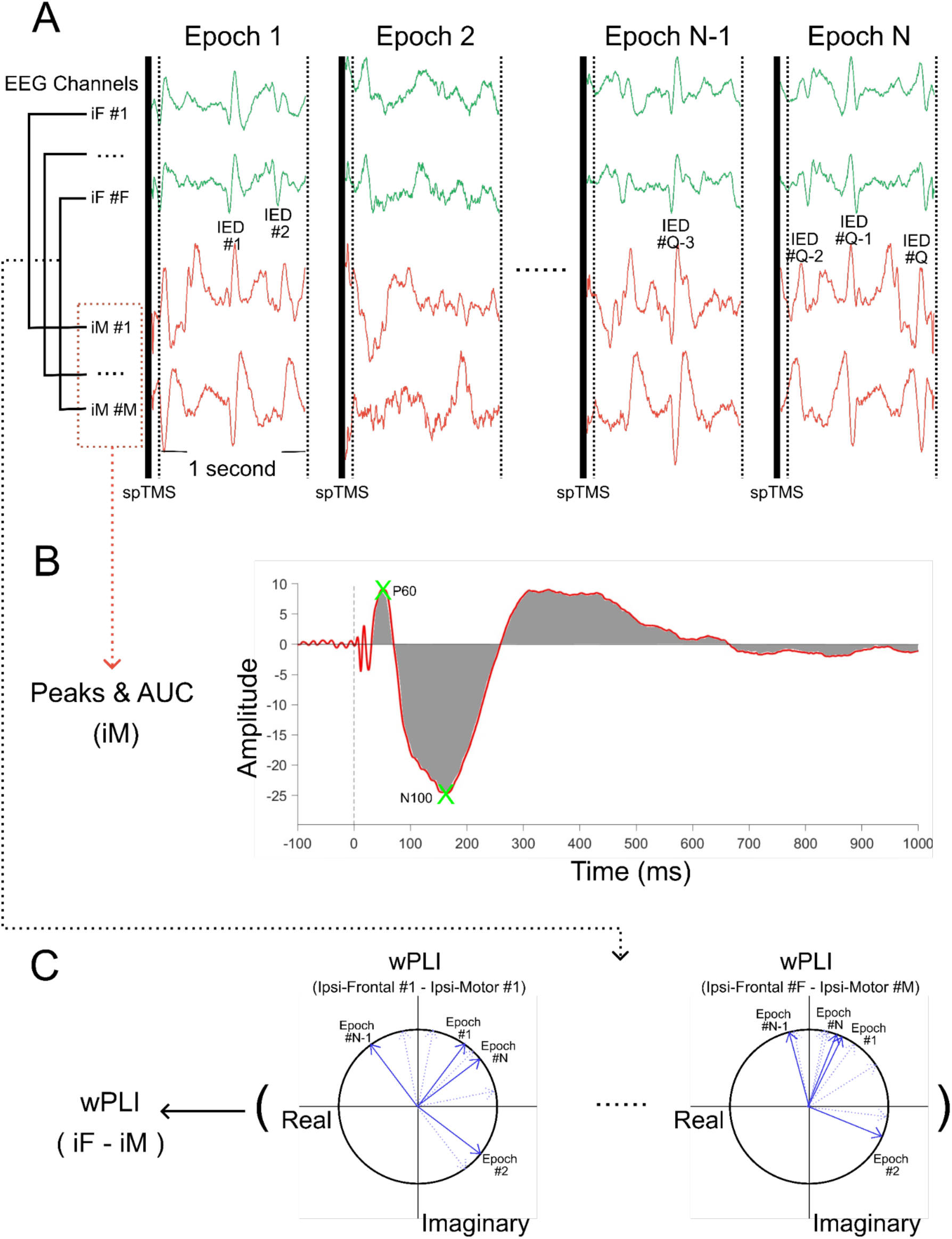
Cortical excitability and connectivity measurement derivation. A: Representative EEG signals measured immediately after single pulses of TMS (spTMS), with exemplary IEDs. One second of EEG is epoched around each spTMS with a matched number of epochs analyzed before and after the rTMS; B: TEP peak amplitudes and latencies as well as area under the curve (AUC) are calculated after averaging all epochs and all channels within a region of interest together. Here we illustrate a TEP from the ipsilateral motor (iM) region; C: Illustration of wPLI calculation between the ipsilateral frontal (iF) and ipsilateral motor (iM) regions.

#### Local TEP

TEPs in children are simpler in morphology compared to adults, and are characterized by a positive peak at 60ms (P60) and a negative peak at 100ms (N100) that can be measured from the stimulated motor cortex (Fig.2B).^22,32,33^ The P60 amplitude was defined as the maximum positive deflection occurring between 30-80 ms, while the N100 was defined as the maximum negative deflection within 80-150 ms following spTMS. The latency of each component was recorded as the time elapsed from spTMS pulse onset to the occurrence of its respective peak.

#### Remote TEPs

We measured the area under the curve (AUC) of both local and remote TEPs at all ROIs. We focused on this measurement as it provides a quantitative measure of the overall signal magnitude and temporal characteristics, and previous reports suggest it changes after rTMS.^34^ The AUC for each TEP was computed by numerically integrating the absolute values of TEP over a predefined time window for each electrode. Then, the AUC value for every electrode within each ROI was averaged to yield a single regional AUC value. Our analyzed time windows are: 1) 10 to 80 millisecond and 2) 10 to 1000 millisecond to encompass the early and full range of TEP characteristics, respectively.^32,33^

#### Weighted phase lag index (wPLI)

We used the wPLI as a second method to assess connectivity between ROIs (Fig. 2C). wPLI is a phase domain metric that is robust against the confounding effects of volume conduction in sensor-based EEG signals.^10^ Beta-band (13–30 Hz) wPLI connectivity was calculated from 10 to 1000 millisecond of TEP using cross-spectral density within MATLAB-based Fieldtrip’s connectivity toolbox implementation. We chose Beta-band because it can be reliably measured across short time periods and has been found to be abnormally elevated in SeLECTS.^4^

First the phase difference between signals from two electrodes (e.g., iF1 and iM1 exemplified in Fig. 2C) was measured based on all time points in the 10-1000 ms window for each epoch. wPLI measures the consistency of phase relationships, assigning greater weight based on the magnitude of the imaginary component, thereby mitigating the impact of volume conduction. The phase differences from all epochs were used to calculate the wPLI between that pair of electrodes. Next, the wPLI value for every electrode pair spanning the two ROIs were averaged to identify a single region-to-region connectivity value:

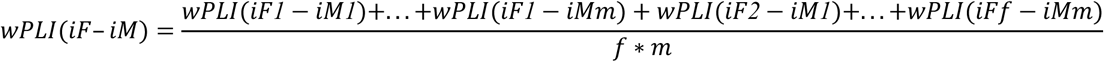

where *iF*=ipsilateral-frontal, *iM*=ipsilateral-motor, 1=first electrode in frontal or motor region, *f*=last electrode in frontal, *m*=last electrode in motor, *f*m*=total number of unique electrode pairs between regions.

#### Frequency of Interictal Epileptiform Discharges (IEDs)

IEDs were manually counted during the blocks of EEG when spTMS was being applied, initially by two research members (XS and WQ) independently and confirmed by a pediatric epileptologist (FMB) who were all blinded to timing (pre vs. post rTMS) or session (sham vs. active rTMS).

### Statistical Analyses

Our primary analyses assessed change of excitability, connectivity, and IED frequency *within* each session. We used paired t-tests to compare the pre- and post-rTMS recordings for the following measurements: (a) amplitude and latency of local TEPs (P60 and N100); (b) AUC of TEPs measured in all six ROIs; (c) wPLI between all six ROIs (15 unique region-to-region pairs); and (d) IED frequency. As a secondary analysis, we compared the change in excitability, connectivity and IED frequency (post minus pre values) induced by active vs. sham rTMS directly with paired t-tests.

Given the multiple ROIs investigated in connectivity analyses, we adjusted our significance threshold using a principal component analysis method, which accounts for the effective number of independent tests.^35^ This method is suitable when data in each comparison are not completely independent, as is the case with EEG where signal represents summated activity from multiple regions.

Previous studies show that connectivity is acutely elevated when IEDs are present.^4^ To quantify the influence of IEDs on connectivity, we tested whether: (a) IED frequency during baseline recordings (pre-rTMS) was associated with baseline connectivity; and (b) change in IED frequency after rTMS was associated with change in connectivity. We modeled these relationships using generalized estimated equations with an exchangeable correlation matrix that can account for repeated measurements.^36^ To understand if rTMS exerted an impact on connectivity beyond its influence on IED, we conducted a sensitivity analysis in which we removed IED-containing epochs and focused on connectivity changes in IED-free epochs.

### Supplementary Analyses

We additionally conducted a series of supplementary analyses. First, as children with epilepsy often have high rMTs exceeding MSO. We thus conducted a sensitivity analysis focusing on participants with measurable rMTs (<84% MSO) to investigate if modulation was more pronounced in these getting appropriate intensity stimulation (Supplementary 1). Next, though we chose beta band a priori as detailed above, we also present changes of connectivity measured in the theta (4–7 Hz) and alpha (8–12 Hz) bands (Supplementary 2) after rTMS. Finally, since both IEDs and connectivity can increase during drowsiness, we closely monitored behavioral state during the experiment to ensure wakefulness. Since power in a variety of frequency bands also changes based on behavioral states,^37^ we analyzed whether evoked power of the theta, alpha, or beta bands changed after rTMS (Supplementary 3).

## Results

### Participants

Twenty-two right-handed children with SeLECTS were enrolled, but three children were excluded: two did not tolerate both sessions and one failed to stay awake. Thus, nineteen children with SeLECTS, aged 7-13 years (10.02±1.44), were included in the analyses. Eleven children (57.9%) were taking ASMs (five took levetiracetam, four took oxcarbazepine, two took both). Average rMT was 89±13% MSO. Average IED frequency was 22+/-42 per 5-minute block; however, six children had the majority of IEDs (>10 IEDs/5-minute), three had rare IEDs (<10 IEDs/5-minute), and remaining children were IED-free during the entire experiment. No medications were changed between experimental sessions.

### Impact of rTMS on Cortical Excitability

#### Local TEP

Amplitude and latency of the P60 and N100 peaks did not change after active or sham rTMS (Fig.3B & Supplementary Table 7).

**Figure 3:**
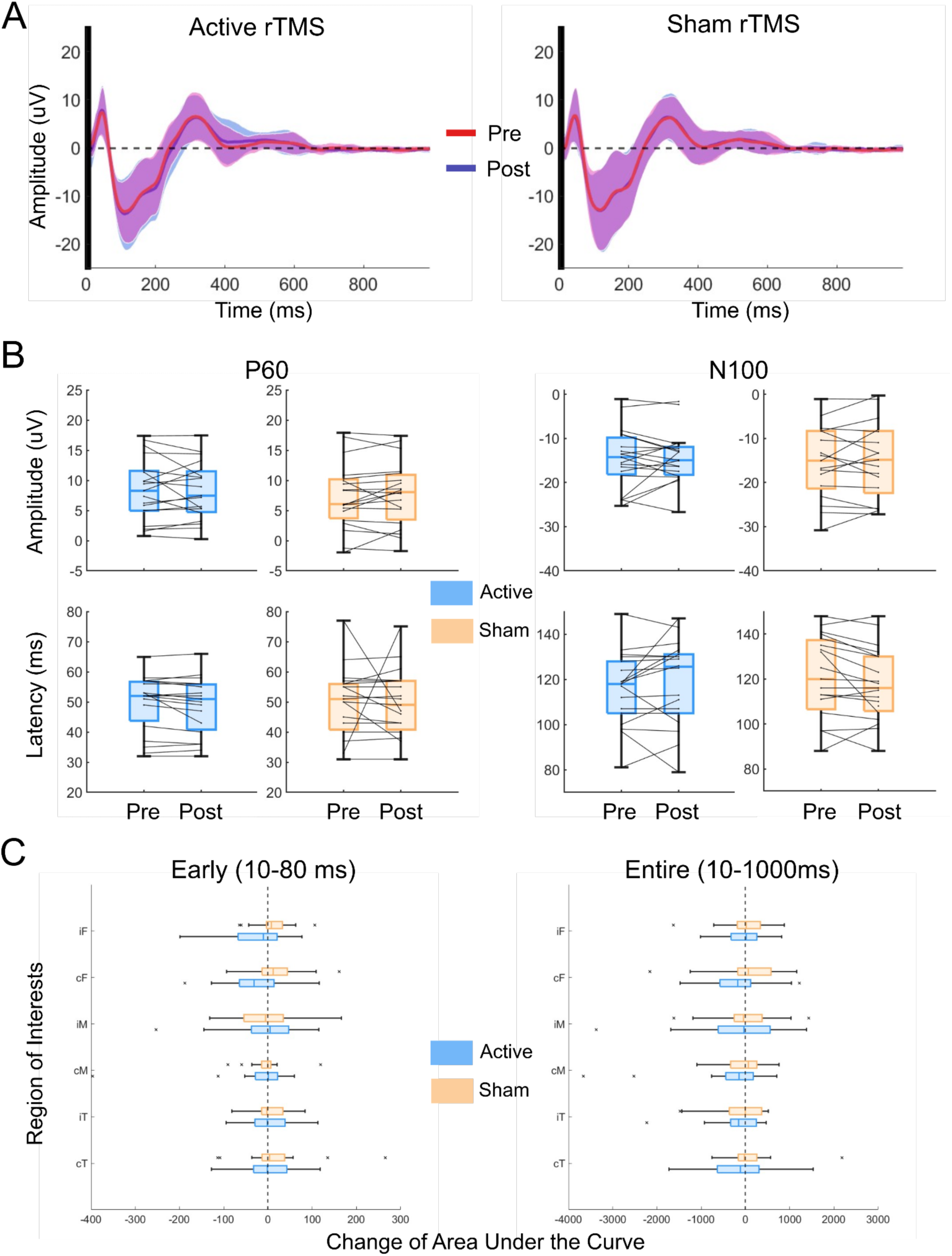
Impact of rTMS on cortical excitability. A: Visualization of the group-average (n=19) local TMS evoked potentials (TEP) at the stimulated site measured before (red) and after (blue) rTMS active (left) and sham (right) rTMS.; B: Change in local TEPs (P60 [left] and N100 [right] amplitudes and latencies) before and after rTMS. There were no significant changes in any of these measurements; C: Change in the area under the curve (AUC) of the TEP in the 10-80 milliseconds (left panel) and 10-1000 milliseconds (right panel) windows for the six regions of interest. There were no significant changes.

### Impact of rTMS on Functional Connectivity

#### Remote TEP

The AUC of the remote TEPs did not change after active or sham rTMS (Fig.3C & Supplementary Table 8) at any ROI.

#### wPLI

In the phase domain, active rTMS significantly reduced wPLI connectivity between seven region-pairs (Fig.4, double asterisks; Supplementary Table 9) whereas no changes were observed after sham rTMS. There was a trend of globally reduced connectivity after active rTMS, though some region-pairs did not meet the adjusted significance threshold (Fig.4, single asterisk). Additionally, in four of the seven region-pairs whose connectivity was significantly reduced compared to baseline, the change also significantly exceeded the change induced by sham rTMS (Fig.4, double daggers).

**Figure 4.**
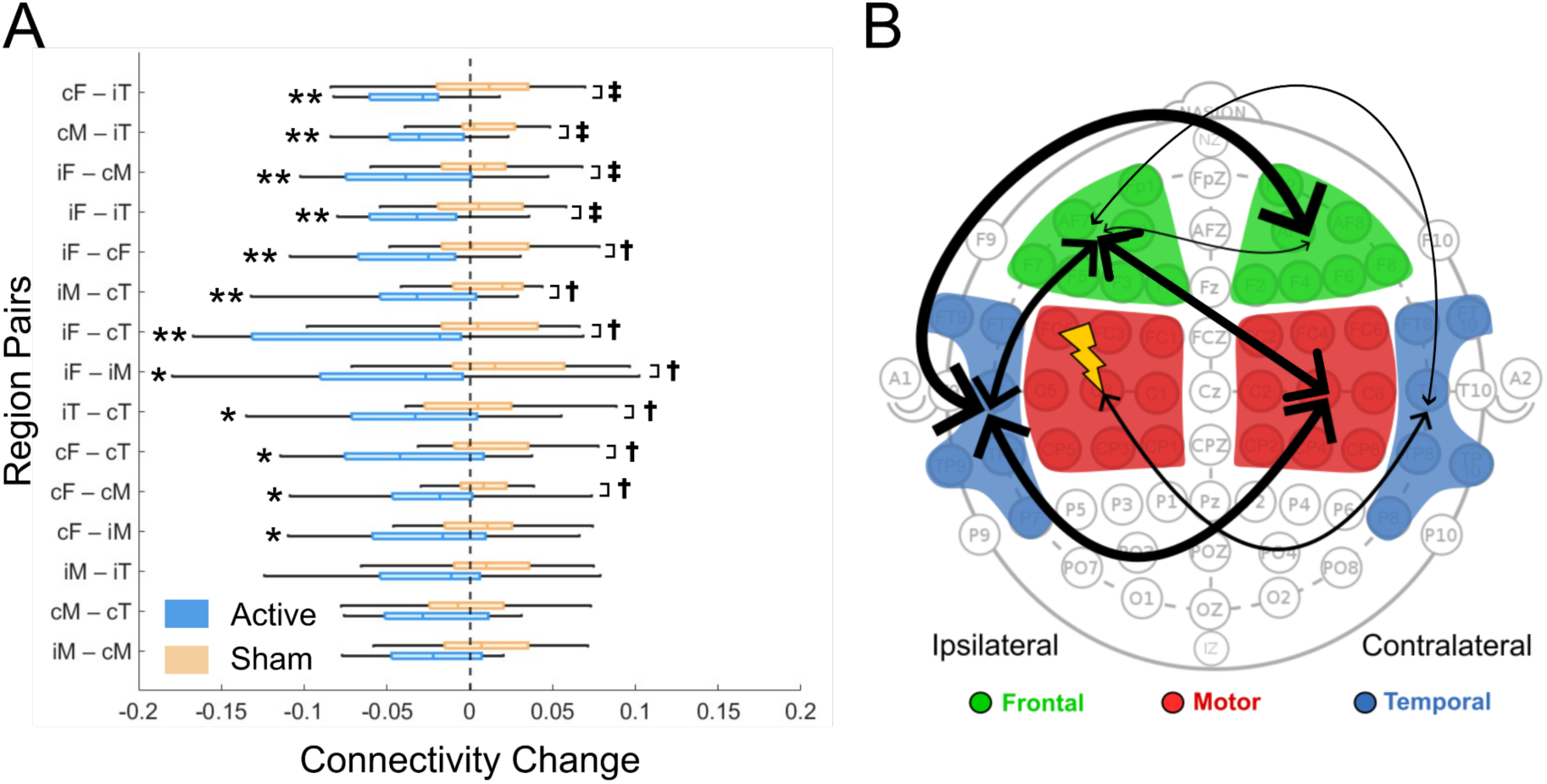
Impact of 1 Hz rTMS on wPLI connectivity. A: Active (blue) but not sham (orange) rTMS reduces connectivity in multiple brain regions in children with SeLECTS. i-: Ipsilateral-, c-: Contralateral-; -F: Frontal, -M: Motor, -T: Temporal. Regions are ordered based on the significance (p values). Dashed line represents no change from baseline. **p<0.0076 (adjusted significance threshold for comparing pre-vs. post-rTMS); ‡p<0.0071 (adjusted significance threshold for comparing active vs. sham rTMS); */† indicate p<0.05 but not meeting the adjusted threshold. B: Black arrows connect regions between which there was a significant reduction in connectivity after active rTMS. Arrow weight based on significance of reduction.

#### Impact of rTMS on IED Frequency

There was no significant change in IED frequency with active or sham rTMS. After active rTMS, IEDs were reduced at a trend level (−11.2±28.8 IEDs/5 mins, p=0.11) whereas after sham rTMS, IED frequency remained more stable (0.8±15.7, p=0.83). The impact of active vs. sham rTMS on IED frequency also did not significantly differ (−12.1±35.3; p=0.16) (Fig.5).

**Figure 5:**
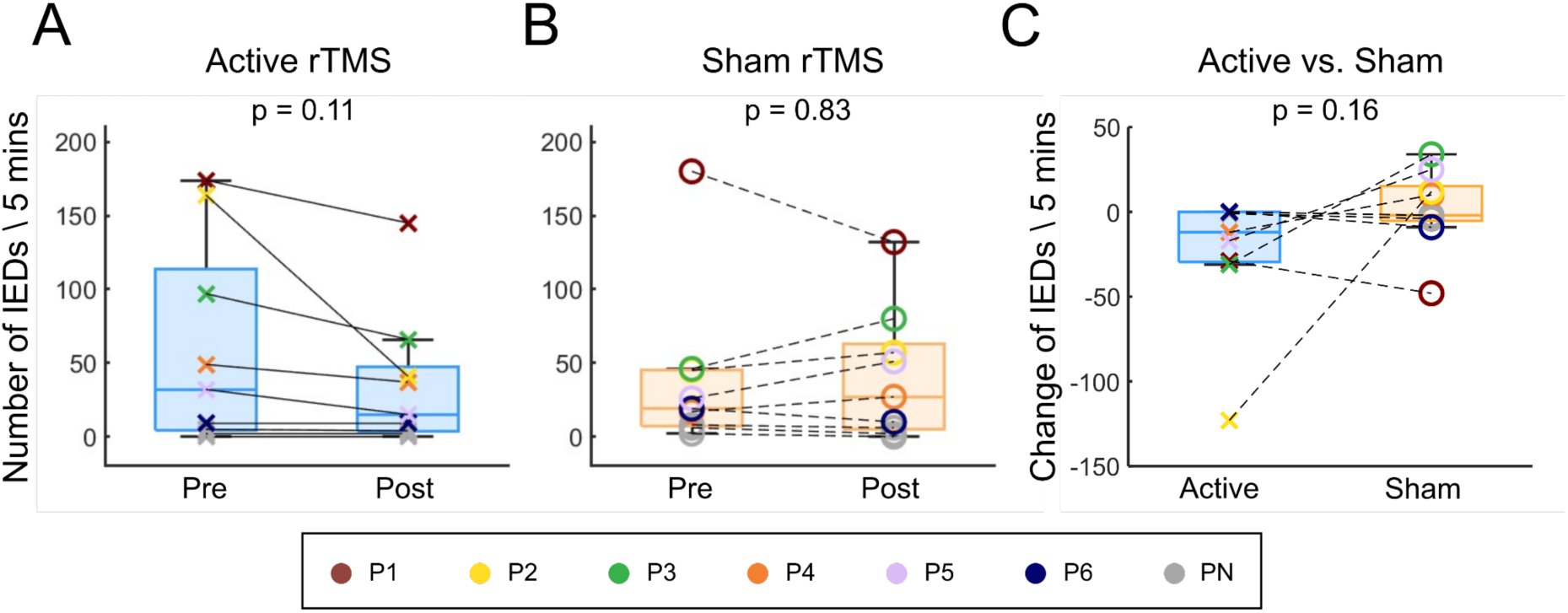
Impact of rTMS on IED frequency in children with SeLECTS. A: Frequency of IEDs pre vs. post active rTMS; B: Frequency of IED pre vs. post sham rTMS; C: change of IED after active vs. sham rTMS. For clarity, each participant with frequent IEDs is represented with an individual color (P1-P6) while the remaining 13 participants with few or no IEDs (PN) are represented in gray.

#### Impact of IEDs on Connectivity

Participants with more IEDs had higher baseline wPLI connectivity (Fig.6A). There was also a significant correlation between change in IED frequency and change in connectivity after both active and sham rTMS (Fig.6B).

**Figure 6:**
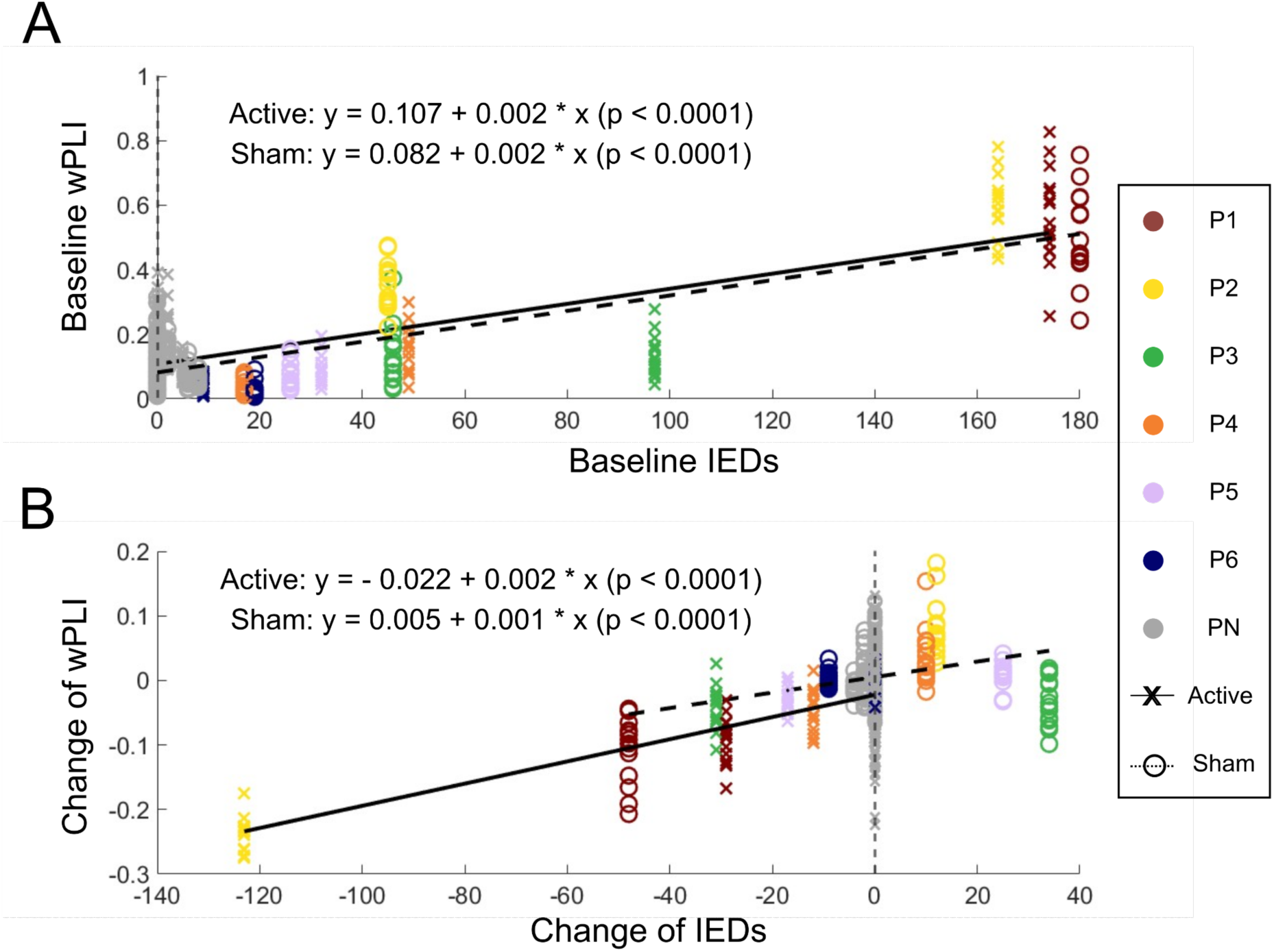
Association of interictal epileptiform discharges (IEDs) and brain connectivity. A: Baseline connectivity measured during both the active (x) and sham (o) sessions was higher in those with higher baseline IED frequency. B: The change in connectivity is significantly associated with change in IED frequency during both the active and sham rTMS sessions. More participants showed a decrease in IEDs on the active day than the sham day. Each x-axis position contains one participant’s connectivity values for all 15 region-pairs. Colored points represent the sub-group of participants (P1-P6) who had frequent IEDs (>10 during baseline recording). Gray points (PN) represent participants who had less than 10 IEDs during the baseline. The solid line is the model fit for the active session and the dashed line is the model fit for the sham session.

#### Impact of rTMS on wPLI Connectivity in IED-free Data

Given that we generally saw a reduction in IEDs after active rTMS and that IEDs themselves were associated with elevated connectivity, we assessed whether the observed reduction in wPLI connectivity (Fig.4) was solely attributable to the decreased IED frequency or could also be measured in IED-free epochs. Results show that reductions in connectivity were also seen in IED-free epochs after active but not sham rTMS (Fig.7 & Supplementary Table 10).

**Figure 7:**
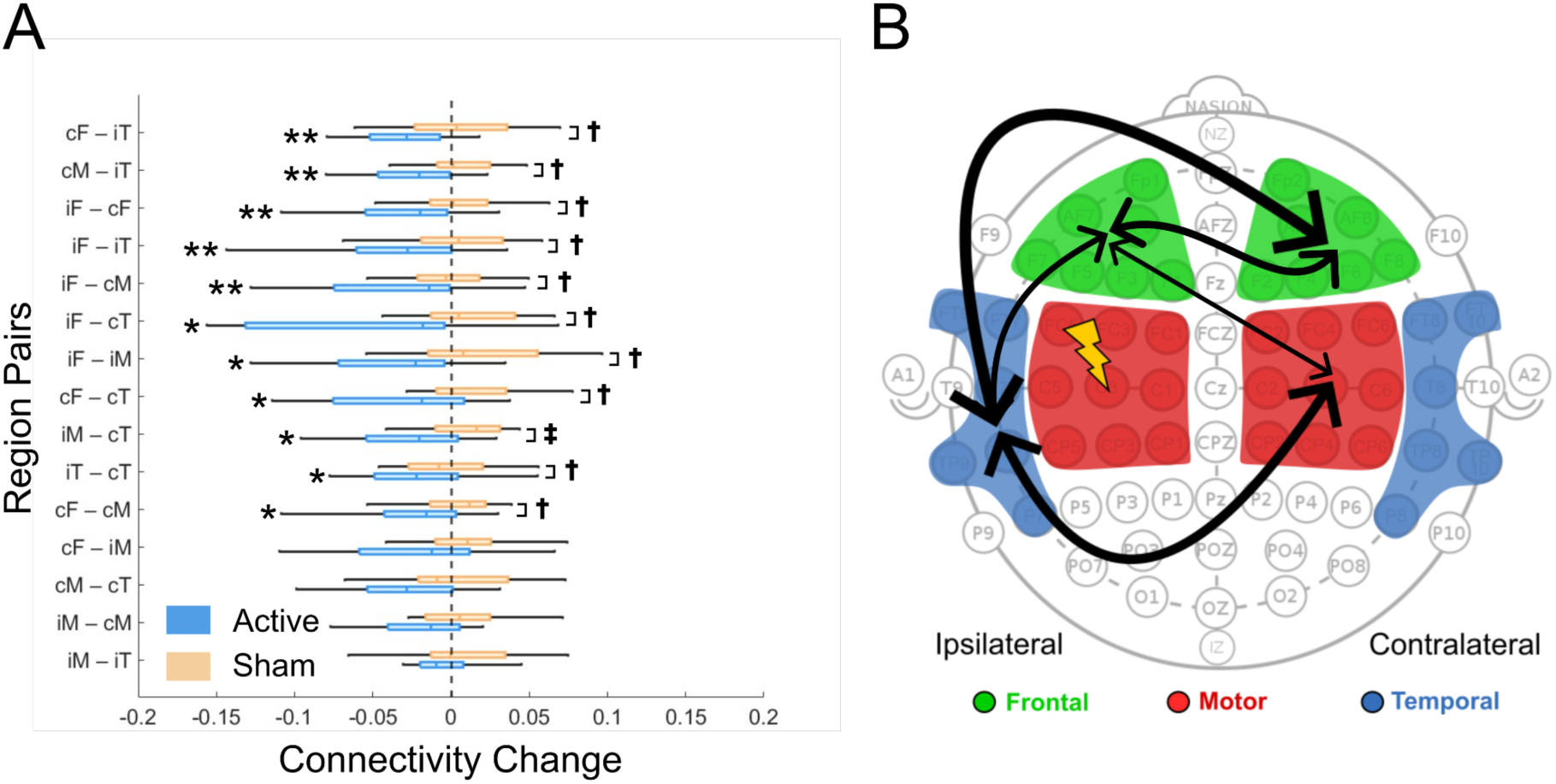
Impact of 1 Hz rTMS on wPLI connectivity during IED-free periods. A: Active (blue) but not sham (orange) rTMS reduces connectivity in multiple brain regions in children with SeLECTS during IED-free period. i-: Ipsilateral-, c-: Contralateral-; -F: Frontal, -M: Motor, -T: Temporal. Regions are ordered based on the significance (p values). Dashed line represents no change from baseline. **p<0.0086 (adjusted significance threshold for comparing pre-vs. post-rTMS); ‡p<0.0091 (adjusted significance threshold for comparing active vs. sham rTMS); */† indicate p<0.05 but not meeting the adjusted threshold. B: Black arrows connect regions between which there was a significant reduction in connectivity after active rTMS. Arrow weight based on significance of reduction.

#### Supplementary Analyses

In a subset of 7 children with lower rMT, we also did not see significant changes in TEP amplitude, latency or AUC (Supplementary 1). In assessing the impact of rTMS on wPLI connectivity in other frequency bands, we found significant reductions across four region-pairs in the theta band after active but not sham rTMS. Additionally, both bands showed broad reductions of connectivity after active rTMS (Supplementary 2). Finally, we found that evoked power did not change with rTMS (Supplementary 3).

## Discussion

Pediatric epilepsy is a severe neurologic disorder that diminishes quality of life not only due to seizures but also because it is associated with neuropsychiatric comorbidities. IEDs, pathologic bursts of hypersynchronous electrical activity, instantaneously disrupt attention and processing speed,^2^ and may lead to long-term alterations in brain functional connectivity.^2–4^ IEDs in childhood furthermore influence brain networks during critical developmental windows. While pharmacologic therapies do not directly target IEDs or connectivity, neurostimulation with rTMS may reduce IED frequency and modulate connectivity in adults.^14,15^ Here, we assessed the impact of active vs. sham rTMS on three neurophysiologic measurements relevant to epilepsy pathology: cortical excitability (measured by TEPs), connectivity (measured using both TEPs and a phase-based measured called wPLI), and IED frequency. We found that rTMS did not affect TEPs. In contrast, rTMS reduced wPLI connectivity widely throughout the brain. Moreover, though limited by a small sample size, the data suggest that rTMS may reduce IEDs.

We investigated the impact of rTMS on cortical excitability because low frequency rTMS has been thought to induce local cortical inhibition, as it suppresses motor evoked potentials (MEPs).^13^ We focused on TEPs instead of MEPs because TEPs capture cortico-cortical interactions rather than only corticomotor ones. Furthermore, TEPs can be reliably measured even in children whose motor threshold exceeds MSO.^6^ Generators of TEPs are not fully understood, but pharmacological studies suggest that the P60 is associated with cortical inhibitory mechanisms,^38^ and the N100 is a long-range inhibitory response related to GABA-B.^8^ Prior studies investigating the effects of low-frequency rTMS on TEPs report mixed findings. Healthy adults showed increases in P60 and N100 amplitude after 1Hz rTMS,^34^ whereas children with untreated attention deficit hyperactivity disorder (ADHD) had reduced N100 amplitudes.^39^ In our pilot study, nine children with SeLECTS showed no significant change in N100 after rTMS on the group level,^22^ and here we similarly found no change in either peak. Several factors may explain discrepancy between studies. One consideration is that children have higher rMT than adults, presumably due to underdeveloped myelination. While it is standard in adults to base TMS intensity on rMT,^40^ this practice may be inappropriate in children, where the disassociation between the intensity needed to elicit an MEP vs. a TEP is wider. Hence, we may be hitting a ceiling effect with TEP amplitudes in children with high rMT such that it is difficult to shift TEPs with rTMS. Arguing against this, we did not see greater TEP modulation when focusing on children with lower rMT (Supplementary 1). A second consideration is that rTMS’s impact may be disease-specific. For instance, children with ADHD might have reduced baseline inhibition and thus be more responsive to inhibitory neuromodulation.^39^ In contrast, children with SeLECTS who have motor cortex pathology may develop compensatory long-range inhibitory input to the motor cortex and thus be less responsive to inhibitory rTMS.^41^ A third consideration is that rTMS may not alter cortical excitability as previously accepted in the literature. Several recent studies failed to find consistent modulation in MEPs or TEPs with several forms of rTMS.^42,43^ The effects of rTMS may be more variable and context-dependent than previously thought, requiring further investigation into disease-specific responses and alternative mechanisms of action.

We next investigated the impact of rTMS on connectivity using two distinct methodologies: a temporal-domain measure (TEPs) and a phase-domain measure (wPLI). The TEP or “inductive” approach,^9^ assumes that connectivity between the stimulated region and remote regions can be quantified by measuring the amplitude of the TEPs in those remote regions, with higher values suggesting stronger connectivity. This method provides temporally specific measurements but is limited to interrogating connectivity of the stimulated cortex. In contrast, wPLI provides inferences about connectivity between regions remote from the site of stimulation.^10^ While TEPs did not change, wPLI connectivity significantly decreased after rTMS, particularly between frontal and temporal regions. Why do inferences from these two measures differ? One possibility is that rTMS influences distributed networks rather than only the stimulated cortex, thus rendering the inductive technique insensitive; with wPLI, we found the largest differences not in motor connectivity but in frontal-to-temporal connectivity. Prior work similarly demonstrated that rTMS of the primary motor cortex (M1) changed TEP amplitudes in bilateral premotor cortices more so than in the stimulated M1.^44^ Another possibility is that the phase-based measures are more sensitive to connectivity changes in epilepsy. In line with this, a study using coherence (another phase-based measure) found an increase in connectivity from baseline in adults with genetic generalized epilepsies but not in controls after TMS pulses.^11^ Since typically-developing controls were not included here for ethical reasons, this prior work contextualizes our findings by suggesting that reduction in connectivity after rTMS indicates normalization of brain responses. Together, these studies highlight the importance of using phase-based measures in addition to evoked potentials to measure the effects of neuromodulation on connectivity.

We next explored whether rTMS modulates IEDs as this is a question of great therapeutic interest. There was a promising trend toward IED reduction, though it was not significant. This aligns with the Cochrane review’s conclusion that there is reasonable evidence that rTMS is effective at reducing IEDs.^14,15^ Our power was limited as only six participants exhibited frequent IEDs during the experiment. This is not unexpected as IEDs are sleep-potentiated in SeLECTS but we measured them during wakefulness, because we wanted to ensure a consistent behavioral state for our excitability and connectivity measurements.^4,45^ Two small open-label studies recently found that multiple doses of 1Hz rTMS reduce IEDs during sleep in children with SeLECTS.^46,47^ Studying the impact of rTMS on IEDs in sleep using rigorous, controlled methods will be a critical next step in determining the therapeutic potential of rTMS for targeting IEDs.

Finally, we assessed if connectivity changes solely reflect IED reduction, as IEDs themselves are associated with short spikes in connectivity.^2,4^ Our results confirm that children with higher IED frequency have higher baseline connectivity and change in IED frequency strongly correlates with change in connectivity. The connectivity reduction was also present in IED-free periods, however, suggesting the effect is not solely due to IED suppression. The directional relationship between IEDs and connectivity remains unclear; rTMS might reduce IEDs, leading to altered connectivity. Alternatively, rTMS may reduce connectivity first, thereby lowering the likelihood of IEDs. Several studies have investigated the relationship between IEDs and connectivity.^48,49^ One found that real IEDs measured from epilepsy patients increased connectivity whereas simulated IEDs introduced into healthy control recordings did not,^49^ suggesting that hyperconnectivity in epilepsy stems from inherent pathophysiological networks rather than just acute effects of IEDs waveform. Another SeLECTS study using causal modeling of EEG-fMRI data showed that IEDs alter network architecture, increasing connectivity between the Rolandic area and the thalamus.^48^ Together, these findings suggest that IEDs may acutely drive hyperconnectivity which then becomes a persistent feature even during periods without IEDs. Although our study did not directly investigate this causality, the observed simultaneous reductions in both IED and connectivity following rTMS highlight the intervention’s therapeutic potential in addressing hyperconnectivity associated with epilepsy.

### Limitations

Certain limitations warrant discussion. First, the spatial specificity of our findings is limited because our analyses were conducted in sensor space due to lack of individual MRIs. While source localization would be valuable, the broad connectivity changes suggest that the impact of rTMS may be fairly distributed rather than isolated to the motor cortex. IEDs in SeLECTS also lead to widespread connectivity increases,^2–4^ suggesting that motor cortex activity (whether spontaneous from IEDs or induced by TMS) influences broad networks. Volume conduction is unlikely to explain these findings as our connectivity metric, wPLI, is stringent against it. A second limitation is that participants did not perform a task confirming equal states of alertness during the measurements, as we did not feel children could do so without compromising EEG quality. TEPs and connectivity can be state-dependent and IEDs are more prominent in SeLECTS during drowsiness and sleep.^4,45^ We mitigated these concerns by monitoring EEG throughout the session and by taking brief breaks after each TMS block to ensure wakefulness. We additionally confirmed that evoked power did not change (Supplementary 3) as power of certain bands varies with drowsiness.^37^ Given these measures, we do not feel that our findings are attributable to state changes. Third, our small sample size limited our ability to account for clinical variables, such as ASMs and age, that may influence the effects of rTMS.^24^ Our crossover design accounts for within-participant variability, but future studies with larger cohorts will be needed. Finally, while this study shows that rTMS reduces connectivity in SeLECTS, it does not establish the clinical utility of doing so. Single rTMS doses are known to induce transient modulation, with repeated doses required to induce durable effects;^50^ here we did not explore the duration of connectivity suppression. Additionally, we did not evaluate the impact of rTMS on cognition, as we generally would not expect measurable changes in cognition within minutes. As prior studies have linked hyperconnectivity with poor outcomes in several pediatric epilepsy syndromes,^5^ however, we believe that further exploration of rTMS as a targeted non-invasive method for modulating connectivity is valuable. Similarly, methods that safely suppress IEDs will permit us to explore their cognitive impact on children with epilepsy. Future studies testing the impact of multi-dose rTMS protocols on functional connectivity, IEDs, and cognitive function are needed.

## Conclusion

This study demonstrates that rTMS can modulate brain connectivity in children with epilepsy and suggests that it may be a non-pharmacological option for reducing IEDs. Together, these findings position rTMS as an important non-invasive method for probing how epilepsy and IEDs affect cognition and suggest that rTMS has therapeutic potential for the treatment of pediatric epilepsy and its cognitive comorbidities.

## Data Availability

All data produced in the present study are available upon reasonable request to the authors

The data analyzed in this study was gathered as part of a larger clinical trial (NCT04325282) of spTMS-EEG in SeLECTS. There are no reproduced materials. The datasets generated and/or analyzed during the current study are not publicly available due to privacy concerns. The data is clinical data that has not been de-identified. But the code with example data used for illustrating the methodologies of TEP and connectivity analysis is publicly available at https://github.com/Pediatric-Neurostimulation-Laboratory/rTMSConnectivity. The full data is available from the corresponding author on reasonable request.

## Acknowledgments

XS receives support from the Stanford Maternal & Child Health Research Institute (MCHRI Postdoctoral Support Fellowship). FMB’s work is supported by a K23 Career Development Award (NINDS K23NS116110), the Doris Duke & Rita Allen Foundation, and a gift from the Principe & O’Farrell family.

## Author Contributions

X.S.: conceptualization, methodology, software, formal analysis, data curation, writing– original draft & editing, visualization. W.Q.: data curation, validation, writing–review & editing. K.C.N.: methodology, software, data curation, validation, writing–review & editing. M.M.: data curation, validation, writing-review & editing. C.C.C.: methodology, software, validation, writing–review & editing. W.W.: methodology, software, writing–review & editing. Z.H.: methodology, writing–review & editing. F.M.B.: conceptualization, methodology, formal analysis, data curation, validation, writing–drafting, review & editing, project administration, funding acquisition.

## References

1. Baumer FM, Cardon AL, Porter BE. Language dysfunction in pediatric epilepsy. J Pediatr. 2018;194:13–21.

2. Holmes GL, Lenck-Santini PP. Role of interictal epileptiform abnormalities in cognitive impairment. Epilepsy Behav. 2006;8(3):504–515.

3. Cheng D, Yan X, Xu K, Zhou X, Chen Q. The effect of interictal epileptiform discharges on cognitive and academic performance in children with idiopathic epilepsy. BMC Neurol. 2020;20:1–7.

4. Goad BS, Lee-Messer C, He Z, Porter BE, Baumer FM. Connectivity increases during spikes and spike-free periods in self-limited epilepsy with centrotemporal spikes. Clin Neurophysiol. 2022;144:123–134.

5. Xiao F, An D, Lei D, et al. Real-time effects of centrotemporal spikes on cognition in rolandic epilepsy: an EEG-fMRI study. Neurology. 2016;86(6):544–551.

6. Tremblay S, Rogasch NC, Premoli I, et al. Clinical utility and prospective of TMS–EEG. Clin Neurophysiol. 2019;130(5):802–844.

7. Kimiskidis V. Transcranial magnetic stimulation (TMS) coupled with electroencephalography (EEG): biomarker of the future. Rev Neurol (Paris). 2016;172(2):123–126.

8. Premoli I, Castellanos N, Rivolta D, et al. TMS-EEG signatures of GABAergic neurotransmission in the human cortex. J Neurosci. 2014;34(16):5603–5612.

9. Bortoletto M, Veniero D, Thut G, Miniussi C. The contribution of TMS–EEG coregistration in the exploration of the human cortical connectome. Neurosci Biobehav Rev. 2015;49:114–124.

10. Vinck M, Oostenveld R, Van Wingerden M, Battaglia F, Pennartz CMA. An improved index of phase-synchronization for electrophysiological data in the presence of volume-conduction, noise and sample-size bias. Neuroimage. 2011;55(4):1548–1565.

11. Vlachos I, Kugiumtzis D, Tsalikakis DG, Kimiskidis VK. TMS-induced brain connectivity modulation in Genetic Generalized Epilepsy. Clin Neurophysiol. 2022;133:83–93.

12. Maeda F, Keenan JP, Tormos JM, Topka H, Pascual-Leone A. Modulation of corticospinal excitability by repetitive transcranial magnetic stimulation. Clin Neurophysiol. 2000;111(5):800–805.

13. Fitzgerald PB, Fountain S, Daskalakis ZJ. A comprehensive review of the effects of rTMS on motor cortical excitability and inhibition. Clin Neurophysiol. 2006;117(12):2584–2596.

14. Walton D, Spencer DC, Nevitt SJ, Michael BD. Transcranial magnetic stimulation for the treatment of epilepsy. Cochrane Database Syst Rev. 2021;(4).

15. Chen R, Spencer DC, Weston J, Nolan SJ. Transcranial magnetic stimulation for the treatment of epilepsy. Cochrane Database Syst Rev. 2016;(8).

16. Specchio N, Wirrell EC, Scheffer IE, et al. International League Against Epilepsy classification and definition of epilepsy syndromes with onset in childhood: Position paper by the ILAE Task Force on Nosology and Definitions. Epilepsia. 2022;63(6):1398–1442.

17. Ostrowski LM, Chinappen DM, Stoyell SM, et al. Children with Rolandic epilepsy have micro-and macrostructural abnormalities in white matter constituting networks necessary for language function. Epilepsy Behav. 2023;144:109254.

18. Rossi S, Antal A, Bestmann S, et al. Safety and recommendations for TMS use in healthy subjects and patient populations, with updates on training, ethical and regulatory issues: Expert Guidelines. Clin Neurophysiol. 2021;132(1):269–306.

19. She X, Nix KC, Cline CC, et al. Stability of transcranial magnetic stimulation electroencephalogram evoked potentials in pediatric epilepsy. Sci Rep. 2024;14(1):9045. doi:10.1038/s41598-024-59468-8

20. Fonov V, Evans AC, Botteron K, et al. Unbiased average age-appropriate atlases for pediatric studies. Neuroimage. 2011;54(1):313–327.

21. Rossini P. Applications of magnetic cortical stimulation. The international federation of clinical neurophysiology. Electroencephalogr Clin Neurophysiol Suppl. 1999;52:171–185.

22. Baumer FM, Pfeifer K, Fogarty A, et al. Cortical Excitability, Synaptic Plasticity & Cognition in Benign Epilepsy with Centrotemporal Spikes: A Pilot TMS-EMG-EEG Study. J Clin Neurophysiol Off Publ Am Electroencephalogr Soc. 2020;37(2):170.

23. Ziemann U, Lönnecker S, Steinhoff BJ, Paulus W. Effects of antiepileptic drugs on motor cortex excitability in humans: a transcranial magnetic stimulation study. Ann Neurol Off J Am Neurol Assoc Child Neurol Soc. 1996;40(3):367–378.

24. Määttä S, Könönen M, Kallioniemi E, et al. Development of cortical motor circuits between childhood and adulthood: A navigated TMS-HdEEG study. Hum Brain Mapp. 2017;38(5):2599–2615.

25. Rosanova M, Casali A, Bellina V, Resta F, Mariotti M, Massimini M. Natural frequencies of human corticothalamic circuits. J Neurosci. 2009;29(24):7679–7685.

26. Delorme A, Makeig S. EEGLAB: an open source toolbox for analysis of single-trial EEG dynamics including independent component analysis. J Neurosci Methods. 2004;134(1):9–21.

27. Cline CC, Lucas MV, Sun Y, Menezes M, Etkin A. Advanced artifact removal for automated TMS-EEG data processing. In: 2021 10th International IEEE/EMBS Conference on Neural Engineering (NER). IEEE; 2021:1039–1042.

28. Mutanen TP, Kukkonen M, Nieminen JO, Stenroos M, Sarvas J, Ilmoniemi RJ. Recovering TMS-evoked EEG responses masked by muscle artifacts. Neuroimage. 2016;139:157–166.

29. Mutanen TP, Metsomaa J, Liljander S, Ilmoniemi RJ. Automatic and robust noise suppression in EEG and MEG: The SOUND algorithm. Neuroimage. 2018;166:135–151.

30. Pion-Tonachini L, Kreutz-Delgado K, Makeig S. ICLabel: An automated electroencephalographic independent component classifier, dataset, and website. NeuroImage. 2019;198:181–197.

31. Rogasch NC, Sullivan C, Thomson RH, et al. Analysing concurrent transcranial magnetic stimulation and electroencephalographic data: A review and introduction to the open-source TESA software. Neuroimage. 2017;147:934–951.

32. Belardinelli P, König F, Liang C, et al. TMS-EEG signatures of glutamatergic neurotransmission in human cortex. Sci Rep. 2021;11(1):8159.

33. Ziemann U. Transcranial magnetic stimulation at the interface with other techniques: a powerful tool for studying the human cortex. The Neuroscientist. 2011;17(4):368–381.

34. Casula EP, Tarantino V, Basso D, et al. Low-frequency rTMS inhibitory effects in the primary motor cortex: Insights from TMS-evoked potentials. Neuroimage. 2014;98:225–232.

35. Galwey NW. A new measure of the effective number of tests, a practical tool for comparing families of non-independent significance tests. Genet Epidemiol Off Publ Int Genet Epidemiol Soc. 2009;33(7):559–568.

36. Zeger SL, Liang KY, Albert PS. Models for longitudinal data: a generalized estimating equation approach. Biometrics. Published online 1988:1049–1060.

37. Bazanova O, Vernon D. Interpreting EEG alpha activity. Neurosci Biobehav Rev. 2014;44:94–110.

38. Rogasch NC, Daskalakis ZJ, Fitzgerald PB. Mechanisms underlying long-interval cortical inhibition in the human motor cortex: a TMS-EEG study. J Neurophysiol. 2013;109(1):89–98.

39. Helfrich C, Pierau SS, Freitag CM, Roeper J, Ziemann U, Bender S. Monitoring cortical excitability during repetitive transcranial magnetic stimulation in children with ADHD: a single-blind, sham-controlled TMS-EEG study. PloS One. 2012;7(11):e50073.

40. Hernandez-Pavon JC, Veniero D, Bergmann TO, et al. TMS combined with EEG: Recommendations and open issues for data collection and analysis. Brain Stimulat. Published online 2023.

41. Englot DJ, Konrad PE, Morgan VL. Regional and global connectivity disturbances in focal epilepsy, related neurocognitive sequelae, and potential mechanistic underpinnings. Epilepsia. 2016;57(10):1546–1557.

42. Magnuson J, Ozdemir MA, Mathieson E, et al. Neuromodulatory effects and reproducibility of the most widely used repetitive transcranial magnetic stimulation protocols. Plos One. 2023;18(6):e0286465.

43. Ozdemir RA, Boucher P, Fried PJ, et al. Reproducibility of cortical response modulation induced by intermittent and continuous theta-burst stimulation of the human motor cortex. Brain Stimulat. 2021;14(4):949–964.

44. Esser S, Huber R, Massimini M, Peterson M, Ferrarelli F, Tononi G. A direct demonstration of cortical LTP in humans: a combined TMS/EEG study. Brain Res Bull. 2006;69(1):86–94.

45. Li Y, Li Y, Sun J, et al. Relationship between brain activity, cognitive function, and sleep spiking activation in new-onset self-limited epilepsy with centrotemporal spikes. Front Neurol. 2022;13:956838.

46. Jin G, Chen J, Du J, et al. Repetitive transcranial magnetic stimulation to treat benign epilepsy with centrotemporal spikes. Brain Stimulat. 2022;15(3):601–604.

47. Yang Y, Han Y, Wang J, et al. Effects of altered excitation–inhibition imbalance by repetitive transcranial magnetic stimulation for self-limited epilepsy with centrotemporal spikes. Front Neurol. 2023;14:1164082.

48. Dai X jian, Yang Y, Wang Y. Interictal epileptiform discharges changed epilepsy-related brain network architecture in BECTS. Brain Imaging Behav. Published online 2022:1–12.

49. Hu DK, Mower A, Shrey DW, Lopour BA. Effect of interictal epileptiform discharges on EEG-based functional connectivity networks. Clin Neurophysiol. 2020;131(5):1087–1098.

50. Thut G, Pascual-Leone A. A review of combined TMS-EEG studies to characterize lasting effects of repetitive TMS and assess their usefulness in cognitive and clinical neuroscience. Brain Topogr. 2010;22:219–232.

