## Supplementart Material 1 for "Repetitive Transcranial Magnetic Stimulation Modulates Brain Connectivity in Children with Self-limited Epilepsy with Centrotemporal Spikes"

**Supplementary Material 1: Effects of rTMS on Cortical Excitability in Participants with Low Resting Motor Threshold**

We conducted a sensitivity analysis to test the effects of 1 Hz rTMS on cortical excitability in a subset of participants whose rMT was low enough that we could administer our spTMS at the planned 120% rMT intensity (rMT< 84%MSO). This supplementary analysis was to test if we were under-stimulating children with high rMT.

**Data**: We included 7 participants whose rMT was lower than 84% MSO at the ipsilateral hemisphere to the rTMS administration. Because we delivered single pulse TMS at the 120% of rMT and rTMS at 90% of rMT, an rMT lower than 84% MSO indicates that the intensity of TMS did not exceed the maximum output of the TMS machine (100% MSO).

**Methods**: For each participant, we tested the same excitability measurements (amplitude and latency of P60 and N100, and AUC) at the stimulated site.

**Results**: As with the excitability results from all participants, rTMS did not exert a significant impact on cortical excitability, as measured by TEP waveform (Supplementary Fig. 1A), or by peak amplitude and latency at the site of stimulation (Supplementary Fig. 1B), or by AUC at the site of stimulation (Supplementary Fig. 1C & Supplementary Table 1).

However, it is worth noting that there is a trend toward decrease in the ipsilateral frontal and contralateral frontal region (p = 0.05), which fits the wPLI reduction shown in the manuscript.

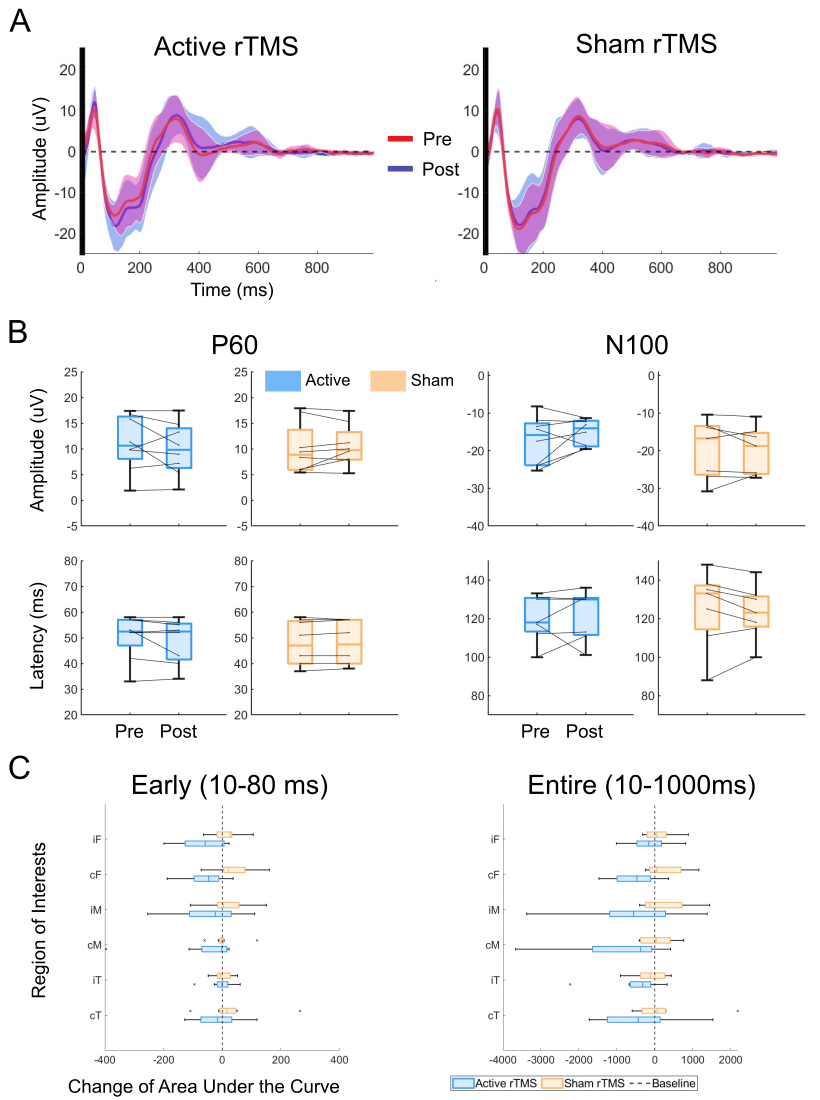

Supplementary Figure 1: change of excitability measurement after active vs. sham rTMS in a subgroup of participants who had rMT lower than 84% MSO.

Supplementary Table 1: Change of AUC after rTMS across participants who had rMT lower than 84% MSO at ROIs. */†p<0.05. i-: Ipsilateral-, c-: Contralateral-; -F: Frontal, -M: Motor, -T: Temporal.

|  | **Active rTMS vs. Baseline** | | | | **Sham rTMS vs. Baseline** | | | | **Active rTMS vs. Sham rTMS** | | | |
| --- | --- | --- | --- | --- | --- | --- | --- | --- | --- | --- | --- | --- |
| **ROI** | **Mean (STD)** | **T** | **p** |  | **Mean (STD)** | **T** | **p** |  | **Mean (STD)** | **T** | **p** |  |
| **Time Window (10-80ms)** | | | | | | | | | | | | |
| **iF** | -66.44 (80.05) | -2.35 | 0.0513 |  | 15.90 (52.06) | 0.86 | 0.4162 |  | -82.34 (109.76) | -2.12 | 0.0715 |  |
| **cF** | -57.46 (70.24) | -2.31 | 0.0539 |  | 37.18 (70.65) | 1.49 | 0.1802 |  | -94.64 (120.62) | -2.22 | 0.0620 |  |
| **iM** | -44.38 (115.08) | -1.09 | 0.3114 |  | 16.64 (76.19) | 0.62 | 0.5562 |  | -61.03 (138.44) | -1.25 | 0.2526 |  |
| **cM** | -60.44 (142.61) | -1.20 | 0.2696 |  | 5.62 (50.28) | 0.32 | 0.7610 |  | -66.06 (147.40) | -1.27 | 0.2454 |  |
| **iT** | -4.05 (45.24) | -0.25 | 0.8076 |  | 1.66 (33.07) | 0.14 | 0.8912 |  | -5.71 (53.89) | -0.30 | 0.7733 |  |
| **cT** | -14.95 (82.38) | -0.51 | 0.6236 |  | 33.29 (106.03) | 0.89 | 0.4040 |  | -48.24 (130.62) | -1.04 | 0.3309 |  |
| **Time Window (10-1000ms)** | | | | | | | | | | | | |
| **iF** | -141.69 (567.4) | -0.71 | 0.5028 |  | 104.78 (394.6) | 0.75 | 0.4771 |  | -246.47 (826.5) | -0.84 | 0.4268 |  |
| **cF** | -533.23 (644.1) | -2.34 | 0.0517 |  | 266.43 (554.5) | 1.36 | 0.2163 |  | -799.65 (905.2) | -2.50 | 0.0411 | † |
| **iM** | -614.78 (1455.9) | -1.19 | 0.2712 |  | 211.43 (679.3) | 0.88 | 0.4079 |  | -826.21 (1562.2) | -1.50 | 0.1783 |  |
| **cM** | -931.35 (1412.7) | -1.86 | 0.1045 |  | 56.52 (449.8) | 0.36 | 0.7328 |  | -987.87 (1513.5) | -1.85 | 0.1074 |  |
| **iT** | -508.87 (765.6) | -1.88 | 0.1022 |  | -122.27 (453.4) | -0.76 | 0.4705 |  | -386.60 (862.3) | -1.27 | 0.2453 |  |
| **cT** | -411.69 (1045.3) | -1.11 | 0.3021 |  | 197.59 (871.5) | 0.64 | 0.5418 |  | -609.28 (1155.7) | -1.49 | 0.1795 |  |
