## Supplementart Material 2 for "Repetitive Transcranial Magnetic Stimulation Modulates Brain Connectivity in Children with Self-limited Epilepsy with Centrotemporal Spikes"

**Supplementary Material 2: Change of wPLI Connectivity in Alpha and Theta Frequency Bands After rTMS**

We conducted an additional analysis to assess if connectivity as measured by wPLI in the theta (4–7 Hz) and alpha (8–12 Hz) frequency bands changed after active or sham 1 Hz rTMS.

**Data**: The same nineteen children with SeLECTS included in the main analysis.

**Methods**: theta- (4–7 Hz) and alpha- (8–12 Hz) band wPLI connectivity was calculated on 10-1000 ms of cleaned TEP data using cross-spectral density within MATLAB-based Fieldtrip’s connectivity toolbox implementation. All preprocessing and parameters were kept the same as the beta-band wPLI method described in the main analysis.

We compared such wPLI before vs. after active or sham rTMS using t-tests. Significant p-value threshold was corrected for multiple comparisons using the same procedure as the main analyses.

**Results**: In the alpha band, wPLI reductions after rTMS were less pronounced compared to those observed in the beta band (Supplementary Fig. 2A). Significant wPLI reductions were observed after active rTMS in contralateral motor to ipsilateral temporal region only. Besides, four region pairs showed a trend of connectivity reduction after active rTMS. In contrast, after sham rTMS, there was no significant change in connectivity between any regions.

Theta band wPLI exhibited a similar global reduction across numerous brain regions following rTMS (Supplementary Fig. 2B). Four region pairs had a significant reduction in wPLI connectivity after active rTMS (contralateral temporal to contralateral motor and ipsilateral temporal, contralateral frontal to ipsilateral frontal and ipsilateral motor). There was also a trend of globally reduced connectivity across most regions. Additionally, consistent with the findings in the beta band, no regions demonstrated significant changes after sham rTMS.

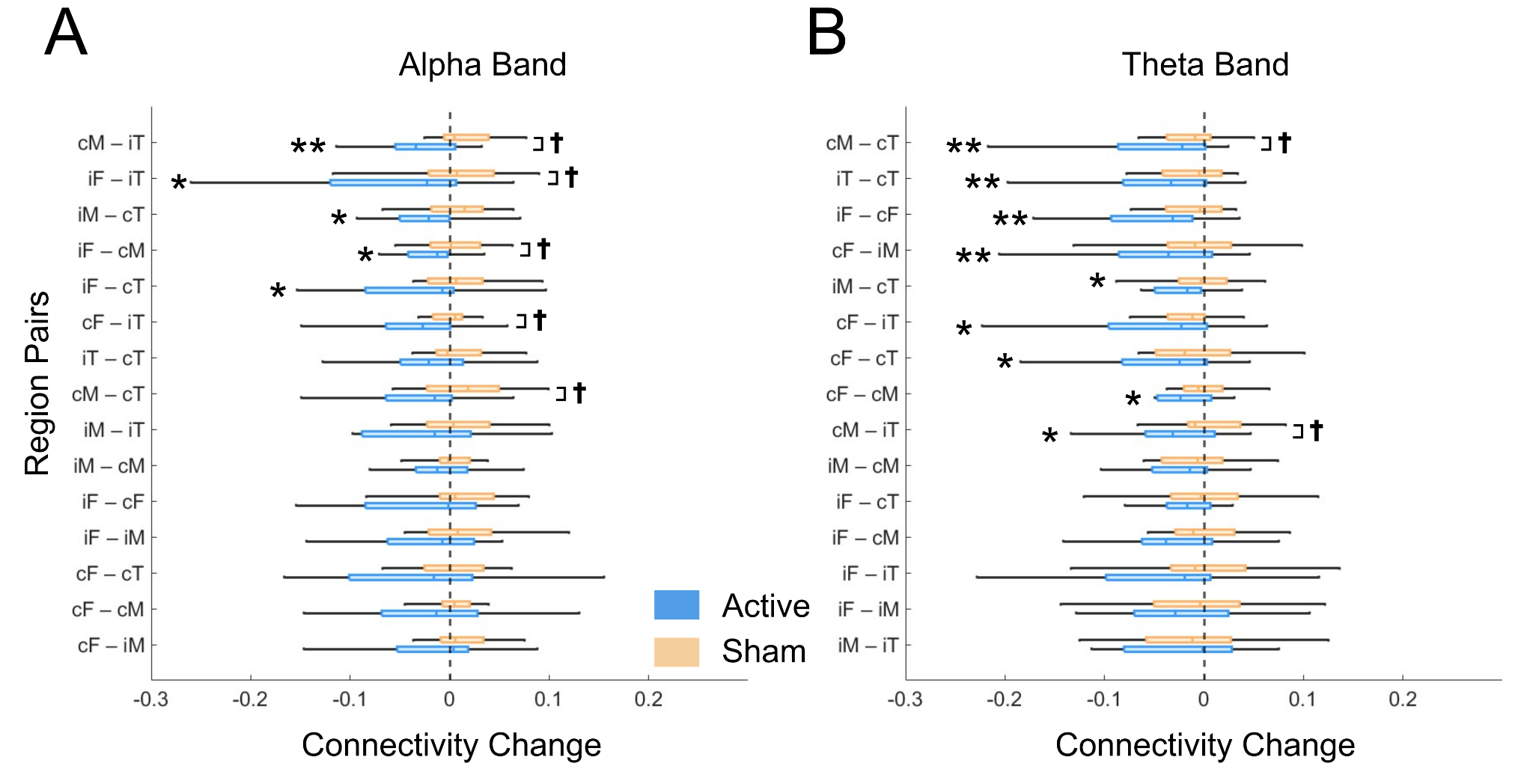

Supplementary Figure 2. Active (blue) but not sham (orange) rTMS reduces wPLI connectivity measured in a) alpha and b) theta frequency bands in multiple brain regions in 19 children with SeLECTS. **p<0.0072 for alpha and **< 0.0107 for theta (adjusted significance threshold in active vs. baseline); */†p<0.05. i-: Ipsilateral-, c-: Contralateral-; -F: Frontal, -M: Motor, -T: Temporal. Regions are ordered based on the significance (p values).

Supplementary Table 2: Change of average **theta band (4-7 Hz)** wPLI connectivity after rTMS across participants at regions of interest pairs. **p<0.0072 (adjusted significance threshold in active vs. baseline); */†p<0.05. i-: Ipsilateral-, c-: Contralateral-; -F: Frontal, -M: Motor, -T: Temporal.

|  | **Active rTMS vs. Baseline** | | | | **Sham rTMS vs. Baseline** | | | | **Active rTMS vs. Sham rTMS** | | | |
| --- | --- | --- | --- | --- | --- | --- | --- | --- | --- | --- | --- | --- |
| **ROI pairs** | **Mean (STD)** | **T** | **p** |  | **Mean (STD)** | **T** | **p** |  | **Mean (STD)** | **T** | **p** |  |
| **iF-cF** | -0.058 (0.081) | -3.04 | 0.0071 | ** | -0.006 (0.061) | -0.42 | 0.6829 |  | -0.052 (0.109) | -2.04 | 0.0561 |  |
| **iF-iM** | -0.054 (0.137) | -1.67 | 0.1128 |  | -0.014 (0.081) | -0.72 | 0.4791 |  | -0.041 (0.148) | -1.17 | 0.2568 |  |
| **iF-cM** | -0.050 (0.110) | -1.95 | 0.0674 |  | 0.007 (0.051) | 0.55 | 0.5917 |  | -0.057 (0.118) | -2.04 | 0.0562 |  |
| **iF-iT** | -0.066 (0.154) | -1.81 | 0.0863 |  | 0.006 (0.075) | 0.31 | 0.7586 |  | -0.071 (0.150) | -2.00 | 0.0606 |  |
| **iF-cT** | -0.041 (0.089) | -1.95 | 0.0672 |  | 0.002 (0.051) | 0.13 | 0.8983 |  | -0.043 (0.106) | -1.70 | 0.1066 |  |
| **cF-iM** | -0.053 (0.078) | -2.88 | 0.0099 | ** | -0.011 (0.052) | -0.93 | 0.3639 |  | -0.041 (0.087) | -2.02 | 0.0589 |  |
| **cF-cM** | -0.053 (0.096) | -2.35 | 0.0302 | * | 0.005 (0.051) | 0.42 | 0.6791 |  | -0.058 (0.121) | -2.03 | 0.0577 |  |
| **cF-iT** | -0.058 (0.099) | -2.48 | 0.0232 | * | -0.011 (0.046) | -1.02 | 0.3204 |  | -0.048 (0.116) | -1.74 | 0.0985 |  |
| **cF-cT** | -0.047 (0.084) | -2.38 | 0.0285 | * | -0.011 (0.049) | -0.93 | 0.3633 |  | -0.037 (0.106) | -1.49 | 0.1536 |  |
| **iM-cM** | -0.038 (0.081) | -1.96 | 0.0657 |  | -0.011 (0.049) | -0.10 | 0.9249 |  | -0.037 (0.086) | -1.82 | 0.0855 |  |
| **iM-iT** | -0.031 (0.085) | -1.54 | 0.1412 |  | -0.012 (0.061) | -0.86 | 0.3997 |  | -0.019 (0.085) | -0.92 | 0.3676 |  |
| **iM-cT** | -0.031 (0.048) | -2.75 | 0.0133 | * | -0.005 (0.038) | -0.56 | 0.5825 |  | -0.027 (0.063) | -1.82 | 0.0853 |  |
| **cM-iT** | -0.046 (0.085) | -2.27 | 0.0357 | * | 0.003 (0.035) | 0.40 | 0.6929 |  | -0.048 (0.091) | -2.22 | 0.0394 | † |
| **cM-cT** | -0.043 (0.060) | -3.07 | 0.0067 | ** | -0.008 (0.038) | -0.95 | 0.3556 |  | -0.035 (0.061) | -2.44 | 0.0255 | † |
| **iT-cT** | -0.044 (0.062) | -3.05 | 0.0070 | ** | -0.015 (0.037) | -1.69 | 0.1076 |  | -0.031 (0.067) | -1.93 | 0.0700 |  |

Supplementary Table 3: Change of average **alpha band (8-12 Hz)** wPLI connectivity after rTMS across participants at regions of interest pairs. **p<0.0107 (adjusted significance threshold in active vs. baseline); */†p<0.05. i-: Ipsilateral-, c-: Contralateral-; -F: Frontal, -M: Motor, -T: Temporal.

|  | **Active rTMS vs. Baseline** | | | | **Sham rTMS vs. Baseline** | | | | **Active rTMS vs. Sham rTMS** | | | |
| --- | --- | --- | --- | --- | --- | --- | --- | --- | --- | --- | --- | --- |
| **ROI pairs** | **Mean (STD)** | **T** | **p** |  | **Mean (STD)** | **T** | **p** |  | **Mean (STD)** | **T** | **p** |  |
| **iF-cF** | -0.031 (0.082) | -1.61 | 0.1252 |  | 0.020 (0.055) | 1.52 | 0.1459 |  | -0.051 (0.114) | -1.89 | 0.0747 |  |
| **iF-iM** | -0.029 (0.079) | -1.58 | 0.1318 |  | 0.018 (0.044) | 1.73 | 0.1016 |  | -0.047 (0.099) | -2.02 | 0.0580 |  |
| **iF-cM** | -0.026 (0.047) | -2.30 | 0.0336 | * | 0.016 (0.068) | 1.01 | 0.3267 |  | -0.042 (0.081) | -2.20 | 0.0409 | † |
| **iF-iT** | -0.054 (0.089) | -2.57 | 0.0191 | * | 0.013 (0.058) | 0.94 | 0.3574 |  | -0.067 (0.127) | -2.26 | 0.0368 | † |
| **iF-cT** | -0.039 (0.074) | -2.26 | 0.0368 | * | 0.017 (0.109) | 0.65 | 0.5262 |  | -0.056 (0.141) | -1.69 | 0.1087 |  |
| **cF-iM** | -0.019 (0.064) | -1.23 | 0.2334 |  | 0.013 (0.043) | 1.26 | 0.2244 |  | -0.031 (0.089) | -1.47 | 0.1584 |  |
| **cF-cM** | -0.025 (0.082) | -1.32 | 0.2045 |  | 0.020 (0.065) | 1.32 | 0.2027 |  | -0.046 (0.100) | -1.94 | 0.0676 |  |
| **cF-iT** | -0.035 (0.074) | -2.05 | 0.0556 |  | 0.013 (0.045) | 1.22 | 0.2382 |  | -0.049 (0.098) | -2.11 | 0.0486 | † |
| **cF-cT** | -0.036 (0.101) | -1.50 | 0.1522 |  | 0.022 (0.121) | 0.78 | 0.4441 |  | -0.058 (0.174) | -1.41 | 0.1755 |  |
| **iM-cM** | -0.021 (0.054) | -1.61 | 0.1243 |  | 0.013 (0.044) | 1.21 | 0.2433 |  | -0.033 (0.074) | -1.89 | 0.0748 |  |
| **iM-iT** | -0.023 (0.057) | -1.67 | 0.1120 |  | 0.012 (0.041) | 1.24 | 0.2304 |  | -0.034 (0.076) | -1.90 | 0.0740 |  |
| **iM-cT** | -0.027 (0.046) | -2.45 | 0.0250 | * | 0.019 (0.064) | 1.26 | 0.2251 |  | -0.045 (0.093) | -2.05 | 0.0554 |  |
| **cM-iT** | -0.030 (0.040) | -3.18 | 0.0051 | ** | 0.019 (0.056) | 1.46 | 0.1608 |  | -0.049 (0.082) | -2.54 | 0.0203 | † |
| **cM-cT** | -0.023 (0.052) | -1.86 | 0.0799 |  | 0.025 (0.067) | 1.57 | 0.1335 |  | -0.048 (0.072) | -2.81 | 0.0117 | † |
| **iT-cT** | -0.029 (0.061) | -2.01 | 0.0595 |  | 0.020 (0.063) | 1.37 | 0.1879 |  | -0.049 (0.111) | -1.88 | 0.0769 |  |
