## Supplementart Material 3 for "Repetitive Transcranial Magnetic Stimulation Modulates Brain Connectivity in Children with Self-limited Epilepsy with Centrotemporal Spikes"

**Supplementary Material 3: Change of Evoked Power after rTMS**

Given the fact that EEG depends on brain states. For example, theta band power increases in drowsiness, whereas posterior alpha power is highest during a quiet, awake state. We conducted an additional analysis to assess if evoked power in the theta (4–7 Hz), alpha (8–12 Hz), and beta (13–30 Hz) bands changed after active or sham 1 Hz rTMS. This supplementary analysis was to examine if the brain states that may be represented by evoked power were altered by rTMS in our cohort.

**Data**: The same nineteen children with SeLECTS included in the main analysis.

**Methods**: We analyzed the event-related spectral perturbation (ERSP) as the measure of evoked power in two time-windows (10–80 ms and 10–1000 ms to the TMS pulse). Specifically, the ERSP was calculated for each electrode by decomposing the spTMS-EEG signal into time-frequency components using the short-time Fourier transform (STFT). This transformation was applied to epochs time-locked to the onset of the TMS pulse spanning over the predefined time windows. The ERSP provides a measure of power changes in various frequency bands over time, relative to the baseline period (-200 to 0 ms). We extracted power values from theta (4–7 Hz), alpha (8–12 Hz), and beta (13–30 Hz) bands by averaging the ERSP values over the selected time window and frequencies. Finally, such power values within the same regions (ipsilateral or contralateral of frontal, motor, or temporal) were averaged.

We compared such evoked power before vs. after active or sham rTMS using t-tests.

**Results**: rTMS did not exert a significant impact on evoked power measured across any frequencies or ROIs in either active or sham condition (Supplementary Fig. 3). Additionally, there was no difference in the impact of active vs. sham rTMS on those measurements.

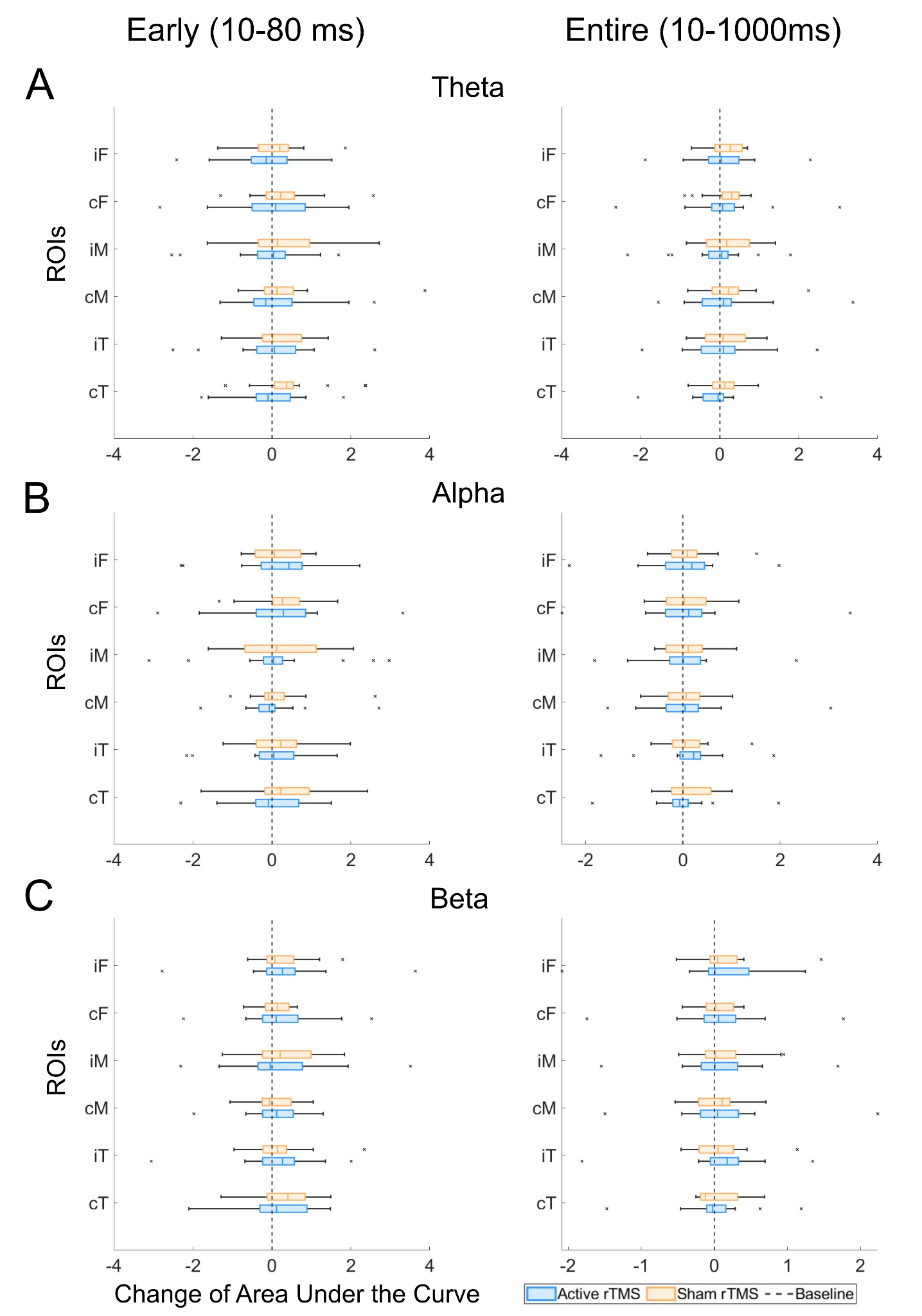

Supplementary Figure 3: change of evoked power in a) theta (4-7 Hz), b) alpha (8-12 Hz), and c) beta (13-30 Hz) frequency band after active vs. sham rTMS. i-: Ipsilateral-, c-: Contralateral-; -F: Frontal, -M: Motor, -T: Temporal.

Supplementary Table 4: Change of theta band (4–7 Hz) evoked power after rTMS across participants at regions of interest. */†p<0.05. i-: Ipsilateral-, c-: Contralateral-; -F: Frontal, -M: Motor, -T: Temporal.

|  | **Active rTMS vs. Baseline** | | | | **Sham rTMS vs. Baseline** | | | | **Active rTMS vs. Sham rTMS** | | |
| --- | --- | --- | --- | --- | --- | --- | --- | --- | --- | --- | --- |
| **ROI** | **Mean (STD)** | **T** | **p** |  | **Mean (STD)** | **T** | **p** |  | **Mean (STD)** | **T** | **p** |
| **Time Window (10-80ms)** | | | | | | | | | | | |
| **iF** | -0.149 (0.935) | 0.68 | 0.5077 |  | 0.125 (0.713) | 0.74 | 0.4688 |  | -0.274 (1.142) | 0.09 | 0.9321 |
| **cF** | 0.038 (1.138) | -0.14 | 0.8884 |  | 0.320 (0.839) | 1.62 | 0.1244 |  | -0.282 (0.839) | -1.00 | 0.3334 |
| **iM** | -0.096 (1.047) | 0.39 | 0.7032 |  | 0.300 (1.076) | 1.18 | 0.2547 |  | -0.395 (1.715) | -0.69 | 0.5001 |
| **cM** | 0.110 (0.955) | -0.49 | 0.6314 |  | 0.335 (0.991) | 1.44 | 0.1693 |  | -0.225 (1.354) | -1.35 | 0.1943 |
| **iT** | -0.034 (1.096) | 0.13 | 0.8961 |  | 0.188 (0.770) | 1.04 | 0.3149 |  | -0.222 (1.518) | -0.58 | 0.5721 |
| **cT** | -0.113 (0.840) | 0.57 | 0.5765 |  | 0.422 (0.906) | 1.98 | 0.0645 |  | -0.535 (1.232) | -1.06 | 0.3044 |
| **Time Window (10-1000ms)** | | | | | | | | | | | |
| **iF** | 0.072 (0.867) | -0.35 | 0.7304 |  | 0.157 (0.453) | 1.47 | 0.1610 |  | -0.085 (0.903) | -0.92 | 0.3687 |
| **cF** | 0.111 (1.070) | -0.44 | 0.6648 |  | 0.194 (0.495) | 1.66 | 0.1147 |  | -0.083 (0.495) | -1.04 | 0.3138 |
| **iM** | -0.096 (0.887) | 0.46 | 0.6534 |  | 0.204 (0.658) | 1.32 | 0.2049 |  | -0.300 (1.181) | -0.45 | 0.6571 |
| **cM** | 0.105 (1.033) | -0.43 | 0.6710 |  | 0.246 (0.676) | 1.54 | 0.1411 |  | -0.141 (1.281) | -1.26 | 0.2259 |
| **iT** | 0.047 (0.939) | -0.21 | 0.8350 |  | 0.132 (0.606) | 0.92 | 0.3680 |  | -0.085 (1.190) | -0.73 | 0.4755 |
| **cT** | -0.085 (0.851) | 0.43 | 0.6756 |  | 0.148 (0.514) | 1.22 | 0.2389 |  | -0.233 (1.124) | -0.31 | 0.7576 |

Supplementary Table 5: Change of alpha band (8–12 Hz) evoked power after rTMS across participants at regions of interest. */†p<0.05. i-: Ipsilateral-, c-: Contralateral-; -F: Frontal, -M: Motor, -T: Temporal.

|  | **Active rTMS vs. Baseline** | | | | **Sham rTMS vs. Baseline** | | | | **Active rTMS vs. Sham rTMS** | | |
| --- | --- | --- | --- | --- | --- | --- | --- | --- | --- | --- | --- |
| **ROI** | **Mean (STD)** | **T** | **p** |  | **Mean (STD)** | **T** | **p** |  | **Mean (STD)** | **T** | **p** |
| **Time Window (10-80ms)** | | | | | | | | | | | |
| **iF** | 0.140 (1.113) | -0.53 | 0.6005 |  | 0.145 (0.628) | 0.98 | 0.3420 |  | -0.005 (1.401) | -1.06 | 0.3045 |
| **cF** | 0.139 (1.311) | -0.45 | 0.659 |  | 0.251 (0.755) | 1.41 | 0.1761 |  | -0.113 (0.755) | -1.21 | 0.2412 |
| **iM** | 0.109 (1.409) | -0.33 | 0.7473 |  | 0.208 (1.044) | 0.85 | 0.4094 |  | -0.099 (2.126) | -1.05 | 0.3071 |
| **cM** | -0.020 (0.875) | 0.10 | 0.9250 |  | 0.155 (0.770) | 0.86 | 0.4041 |  | -0.175 (1.303) | -0.57 | 0.5762 |
| **iT** | 0.030 (0.958) | -0.13 | 0.8961 |  | 0.221 (0.771) | 1.22 | 0.2404 |  | -0.191 (1.536) | -1.31 | 0.2090 |
| **cT** | -0.027 (0.898) | 0.13 | 0.8992 |  | 0.344 (1.020) | 1.43 | 0.1708 |  | -0.371 (1.541) | -1.17 | 0.2581 |
| **Time Window (10-1000ms)** | | | | | | | | | | | |
| **iF** | 0.025 (0.850) | -0.12 | 0.9022 |  | 0.083 (0.518) | 0.68 | 0.5057 |  | -0.058 (1.077) | -0.51 | 0.6198 |
| **cF** | 0.093 (1.107) | -0.36 | 0.7250 |  | 0.071 (0.561) | 0.54 | 0.5975 |  | 0.022 (0.561) | -0.62 | 0.5419 |
| **iM** | -0.006 (0.818) | 0.03 | 0.9753 |  | 0.089 (0.478) | 0.79 | 0.4404 |  | -0.095 (1.083) | -0.45 | 0.6610 |
| **cM** | 0.095 (0.928) | -0.44 | 0.6684 |  | 0.065 (0.523) | 0.53 | 0.6051 |  | 0.030 (1.213) | -0.76 | 0.4570 |
| **iT** | 0.147 (0.713) | -0.87 | 0.3945 |  | 0.084 (0.484) | 0.73 | 0.4728 |  | 0.063 (0.956) | -1.29 | 0.2131 |
| **cT** | -0.037 (0.712) | 0.22 | 0.8287 |  | 0.125 (0.481) | 1.10 | 0.2858 |  | -0.162 (0.988) | -0.53 | 0.6045 |

Supplementary Table 6: Change of beta band (13–30 Hz) evoked power after rTMS across participants at regions of interest. */†p<0.05. i-: Ipsilateral-, c-: Contralateral-; -F: Frontal, -M: Motor, -T: Temporal.

|  | **Active rTMS vs. Baseline** | | | | **Sham rTMS vs. Baseline** | | | | **Active rTMS vs. Sham rTMS** | | |
| --- | --- | --- | --- | --- | --- | --- | --- | --- | --- | --- | --- |
| **ROI** | **Mean (STD)** | **T** | **p** |  | **Mean (STD)** | **T** | **p** |  | **Mean (STD)** | **T** | **p** |
| **Time Window (10-80ms)** | | | | | | | | | | | |
| **iF** | 0.320 (1.200) | -1.13 | 0.2741 |  | 0.250 (0.593) | 1.79 | 0.0909 |  | 0.069 (1.238) | -1.69 | 0.1094 |
| **cF** | 0.278 (1.077) | -1.10 | 0.2886 |  | 0.101 (0.368) | 1.16 | 0.2614 |  | 0.177 (0.368) | -1.52 | 0.1458 |
| **iM** | 0.215 (1.248) | -0.73 | 0.4752 |  | 0.289 (0.927) | 1.32 | 0.2035 |  | -0.074 (1.686) | -1.51 | 0.1483 |
| **cM** | 0.142 (0.741) | -0.81 | 0.4283 |  | 0.056 (0.544) | 0.43 | 0.6693 |  | 0.086 (1.051) | -1.10 | 0.2886 |
| **iT** | 0.184 (1.058) | -0.74 | 0.4712 |  | 0.182 (0.767) | 1.01 | 0.328 |  | 0.002 (1.549) | -1.54 | 0.1425 |
| **cT** | 0.181 (0.902) | -0.85 | 0.4060 |  | 0.285 (0.733) | 1.65 | 0.1168 |  | -0.104 (1.164) | -1.70 | 0.1065 |
| **Time Window (10-1000ms)** | | | | | | | | | | | |
| **iF** | 0.059 (0.675) | -0.37 | 0.7131 |  | 0.116 (0.409) | 1.21 | 0.2435 |  | -0.057 (0.742) | -0.90 | 0.3829 |
| **cF** | 0.074 (0.681) | -0.46 | 0.6499 |  | 0.047 (0.230) | 0.86 | 0.4007 |  | 0.028 (0.230) | -0.75 | 0.4656 |
| **iM** | 0.062 (0.618) | -0.42 | 0.6776 |  | 0.122 (0.407) | 1.28 | 0.2190 |  | -0.061 (0.805) | -1.17 | 0.2590 |
| **cM** | 0.076 (0.720) | -0.45 | 0.6581 |  | 0.062 (0.346) | 0.76 | 0.4550 |  | 0.014 (0.902) | -0.87 | 0.3988 |
| **iT** | 0.117 (0.601) | -0.83 | 0.4193 |  | 0.086 (0.369) | 0.99 | 0.3354 |  | 0.031 (0.769) | -1.36 | 0.1920 |
| **cT** | 0.016 (0.513) | -0.13 | 0.8974 |  | 0.076 (0.339) | 0.96 | 0.3527 |  | -0.060 (0.698) | -0.75 | 0.4613 |
