## Supplementart Table 7 for "Repetitive Transcranial Magnetic Stimulation Modulates Brain Connectivity in Children with Self-limited Epilepsy with Centrotemporal Spikes"

Supplementary Table 7: Change of peak amplitude and latency after rTMS across participants at the stimulated motor cortex.

|  | **Active rTMS vs. Baseline** | | | | **Sham rTMS vs. Baseline** | | | | **Active rTMS vs. Sham rTMS** | | |
| --- | --- | --- | --- | --- | --- | --- | --- | --- | --- | --- | --- |
|  | **Mean (STD)** | **T** | **p** |  | **Mean (STD)** | **T** | **p** |  | **Mean (STD)** | **T** | **p** |
| **P60 Amp.** | -0.25 (2.42) | -0.46 | 0.65 |  | 0.34  (2.09) | 0.72 | 0.48 |  | -0.60 (3.48) | -0.75 | 0.47 |
| **P60 Lat.** | -0.89 (0.58) | -1.51 | 0.15 |  | -0.32 (12.80) | -0.11 | 0.92 |  | -0.58 (12.74) | -0.20 | 0.85 |
| **N100 Amp.** | 0.26  (3.95) | 0.29 | 0.78 |  | -0.72 (5.39) | -0.58 | 0.57 |  | 0.98  (7.45) | 0.57 | 0.57 |
| **N100 Lat.** | 2.67 (10.79) | 1.05 | 0.31 |  | -0.32 (17.63) | -0.08 | 0.94 |  | 2.44 (19.73) | 0.59 | 0.61 |
