## Supplementart Table 8 for "Repetitive Transcranial Magnetic Stimulation Modulates Brain Connectivity in Children with Self-limited Epilepsy with Centrotemporal Spikes"

Supplementary Table 8: Change of AUC after rTMS across participants at regions of interest.

|  | **Active rTMS vs. Baseline** | | | | **Sham rTMS vs. Baseline** | | | | **Active rTMS vs. Sham rTMS** | | |
| --- | --- | --- | --- | --- | --- | --- | --- | --- | --- | --- | --- |
| **ROI** | **Mean (STD)** | **T** | **p** |  | **Mean (STD)** | **T** | **p** |  | **Mean (STD)** | **T** | **p** |
| **Time Window (10-80ms)** | | | | | | | | | | | |
| **iF** | -28.58 (70.55) | -1.77 | 0.0944 |  | 11.58 (41.92) | 1.20 | 0.2442 |  | -40.15 (91.02) | -1.92 | 0.0704 |
| **cF** | -30.55 (67.51) | -1.97 | 0.0641 |  | 18.27 (58.79) | 1.35 | 0.1922 |  | -48.83 (102.75) | -2.07 | 0.0530 |
| **iM** | -11.68 (89.37) | -0.57 | 0.5758 |  | 0.93 (84.37) | 0.05 | 0.9622 |  | -12.62 (132.07) | -0.42 | 0.6821 |
| **cM** | -23.98 (98.31) | -1.06 | 0.3018 |  | -3.61 (39.89) | -0.39 | 0.6978 |  | -20.37 (105.66) | -0.84 | 0.4118 |
| **iT** | 6.47 (52.08) | 0.54 | 0.5948 |  | 3.52 (38.23) | 0.40 | 0.6928 |  | 2.95 (62.35) | 0.21 | 0.8390 |
| **cT** | 0.96 (62.89) | 0.07 | 0.9477 |  | 16.60 (81.75) | 0.89 | 0.3877 |  | -15.64 (105.05) | -0.65 | 0.5245 |
| **Time Window (10-1000ms)** | | | | | | | | | | | |
| **iF** | -6.43 (495.2) | -0.06 | 0.9555 |  | 19.13 (559.8) | 0.15 | 0.8833 |  | -25.56 (836.7) | -0.13 | 0.8956 |
| **cF** | -190.63 (716.0) | -1.16 | 0.2610 |  | 50.85 (793.3) | 0.28 | 0.7831 |  | -241.48 (1201.0) | -0.88 | 0.3923 |
| **iM** | -165.89 (1070.5) | -0.68 | 0.5080 |  | 20.49 (708.7) | 0.13 | 0.9011 |  | -186.38 (1407.6) | -0.58 | 0.5710 |
| **cM** | -369.23 (1038.2) | -1.55 | 0.1385 |  | -10.37 (446.3) | -0.10 | 0.9205 |  | -358.86 (1160.8) | -1.35 | 0.1945 |
| **iT** | -200.59 (622.9) | -1.40 | 0.1774 |  | -127.94 (600.9) | -0.93 | 0.3657 |  | -72.65 (775.4) | -0.41 | 0.6878 |
| **cT** | -192.82 (782.1) | -1.07 | 0.2967 |  | 54.17 (620.0) | 0.38 | 0.7078 |  | -246.99 (941.0) | -1.14 | 0.2676 |
