## Supplementart Table 9 for "Repetitive Transcranial Magnetic Stimulation Modulates Brain Connectivity in Children with Self-limited Epilepsy with Centrotemporal Spikes"

Supplementary Table 9: Change of average wPLI connectivity after rTMS across participants at regions of interest pairs. **p<0.0076 (adjusted significance threshold in active vs. baseline); ‡p<0.0071 (adjusted significance threshold in active vs. sham); */†p<0.05. i-: Ipsilateral-, c-: Contralateral-; -F: Frontal, -M: Motor, -T: Temporal.

|  | **Active rTMS vs. Baseline** | | | | **Sham rTMS vs. Baseline** | | | | **Active rTMS vs. Sham rTMS** | | | |
| --- | --- | --- | --- | --- | --- | --- | --- | --- | --- | --- | --- | --- |
| **ROI pairs** | **Mean (STD)** | **T** | **p** |  | **Mean (STD)** | **T** | **p** |  | **Mean (STD)** | **T** | **p** |  |
| **iF-cF** | -0.033 (0.041) | -3.46 | 0.0028 | ** | 0.009 (0.038) | 1.08 | 0.2927 |  | -0.043 (0.062) | -3.03 | 0.0073 | † |
| **iF-iM** | -0.042 (0.065) | -2.81 | 0.0115 | * | 0.011 (0.065) | 0.75 | 0.4609 |  | -0.053 (0.088) | -2.61 | 0.0176 | † |
| **iF-cM** | -0.037 (0.043) | -3.75 | 0.0015 | ** | 0.007 (0.049) | 0.62 | 0.5445 |  | -0.045 (0.061) | -3.20 | 0.0050 | ‡ |
| **iF-iT** | -0.044 (0.052) | -3.69 | 0.0017 | ** | 0.011 (0.048) | 1.03 | 0.3154 |  | -0.056 (0.079) | -3.11 | 0.0060 | ‡ |
| **iF-cT** | -0.053 (0.073) | -3.18 | 0.0052 | ** | 0.003 (0.076) | 0.16 | 0.8747 |  | -0.056 (0.104) | -2.35 | 0.0304 | † |
| **cF-iM** | -0.022 (0.044) | -2.15 | 0.0456 | * | 0.005 (0.039) | 0.55 | 0.5914 |  | -0.026 (0.065) | -1.70 | 0.1057 |  |
| **cF-cM** | -0.027 (0.050) | -2.40 | 0.0275 | * | 0.012 (0.040) | 1.33 | 0.2005 |  | -0.040 (0.071) | -2.44 | 0.0255 | † |
| **cF-iT** | -0.038 (0.034) | -4.91 | 0.0001 | ** | 0.004 (0.040) | 0.40 | 0.6925 |  | -0.042 (0.043) | -4.29 | 0.0004 | ‡ |
| **cF-cT** | -0.042 (0.066) | -2.77 | 0.0127 | * | 0.014 (0.058) | 1.05 | 0.3056 |  | -0.055 (0.092) | -2.62 | 0.0175 | † |
| **iM-cM** | -0.020 (0.053) | -1.66 | 0.1152 |  | 0.005 (0.040) | 0.54 | 0.5960 |  | -0.025 (0.060) | -1.84 | 0.0817 |  |
| **iM-iT** | -0.026 (0.065) | -1.72 | 0.1017 |  | 0.006 (0.043) | 0.57 | 0.5734 |  | -0.032 (0.073) | -1.91 | 0.0726 |  |
| **iM-cT** | -0.031 (0.040) | -3.43 | 0.0030 | ** | 0.006 (0.056) | 0.45 | 0.6576 |  | -0.037 (0.058) | -2.78 | 0.0123 | † |
| **cM-iT** | -0.029 (0.030) | -4.25 | 0.0005 | ** | 0.006 (0.034) | 0.81 | 0.4273 |  | -0.035 (0.036) | -4.20 | 0.0005 | ‡ |
| **cM-cT** | -0.017 (0.045) | -1.66 | 0.1152 |  | 0.004 (0.044) | 0.41 | 0.6864 |  | -0.022 (0.059) | -1.58 | 0.1326 |  |
| **iT-cT** | -0.033 (0.052) | -2.80 | 0.0119 | * | -0.000 (0.050) | -0.01 | 0.9862 |  | -0.033 (0.062) | -2.35 | 0.0303 | † |
