## Supplementart Table 10 for "Repetitive Transcranial Magnetic Stimulation Modulates Brain Connectivity in Children with Self-limited Epilepsy with Centrotemporal Spikes"

Supplementary Table 10: Change of average wPLI connectivity after rTMS across participants at regions of interest pairs in IED-free data. **p<0.0086 (adjusted significance threshold in active vs. baseline); ‡p<0.0091 (adjusted significance threshold in active vs. sham); */†p<0.05.

|  | **Active rTMS vs. Baseline** | | | | **Sham rTMS vs. Baseline** | | | | **Active rTMS vs. Sham rTMS** | | | |
| --- | --- | --- | --- | --- | --- | --- | --- | --- | --- | --- | --- | --- |
| **ROI pairs** | **Mean (STD)** | **T** | **p** |  | **Mean (STD)** | **T** | **p** |  | **Mean (STD)** | **T** | **p** |  |
| **iF-cF** | -0.030 (0.040) | -3.22 | 0.0047 | ** | 0.013 (0.050) | 1.15 | 0.2662 |  | -0.044 (0.072) | -2.65 | 0.0164 | † |
| **iF-iM** | -0.042 (0.069) | -2.62 | 0.0172 | * | 0.025 (0.063) | 1.75 | 0.0978 |  | -0.066 (0.103) | -2.80 | 0.0117 | † |
| **iF-cM** | -0.033 (0.047) | -3.10 | 0.0062 | ** | 0.013 (0.058) | 0.94 | 0.3596 |  | -0.046 (0.079) | -2.55 | 0.0199 | † |
| **iF-iT** | -0.040 (0.056) | -3.14 | 0.0057 | ** | 0.013 (0.062) | 0.89 | 0.3870 |  | -0.054 (0.110) | -2.13 | 0.0468 | † |
| **iF-cT** | -0.061 (0.101) | -2.63 | 0.0168 | * | 0.013 (0.096) | 0.60 | 0.5554 |  | -0.075 (0.119) | -2.73 | 0.0138 | † |
| **cF-iM** | -0.022 (0.046) | -2.09 | 0.0507 |  | 0.019 (0.054) | 1.53 | 0.1428 |  | -0.040 (0.084) | -2.08 | 0.0518 |  |
| **cF-cM** | -0.025 (0.049) | -2.18 | 0.0430 | * | 0.020 (0.061) | 1.42 | 0.1729 |  | -0.045 (0.086) | -2.25 | 0.0373 | † |
| **cF-iT** | -0.033 (0.034) | -4.27 | 0.0005 | ** | 0.013 (0.064) | 0.86 | 0.3986 |  | -0.046 (0.075) | -2.69 | 0.0149 | † |
| **cF-cT** | -0.046 (0.081) | -2.49 | 0.0227 | * | 0.023 (0.080) | 1.28 | 0.2153 |  | -0.069 (0.111) | -2.71 | 0.0143 | † |
| **iM-cM** | -0.017 (0.052) | -1.42 | 0.1723 |  | 0.013 (0.045) | 1.29 | 0.2125 |  | -0.030 (0.072) | -1.82 | 0.0847 |  |
| **iM-iT** | -0.021 (0.068) | -1.37 | 0.1861 |  | 0.013 (0.052) | 1.06 | 0.3015 |  | -0.035 (0.089) | -1.71 | 0.1045 |  |
| **iM-cT** | -0.031 (0.055) | -2.48 | 0.0232 | * | 0.012 (0.051) | 1.06 | 0.3049 |  | -0.043 (0.064) | -2.94 | 0.0088 | ‡ |
| **cM-iT** | -0.024 (0.029) | -3.57 | 0.0022 | ** | 0.008 (0.051) | 0.66 | 0.5160 |  | -0.031 (0.055) | -2.48 | 0.0234 | † |
| **cM-cT** | -0.020 (0.047) | -1.89 | 0.0756 |  | 0.008 (0.048) | 0.69 | 0.4960 |  | -0.028 (0.066) | -1.89 | 0.0757 |  |
| **iT-cT** | -0.033 (0.062) | -2.31 | 0.0331 | * | -0.000 (0.052) | -0.07 | 0.9442 |  | -0.032 (0.064) | -2.18 | 0.0429 | † |
